# Trajectories of gene expression, seasonal influenza, and within-host seasonal immunity: transfer value to covid-19

**DOI:** 10.1101/2022.03.01.22271679

**Authors:** Eiliv Lund, Marit Holden, Lill-Tove Rasmussen Busund, Igor Snapkov, Nikita Shvetsov, Lars Holden

## Abstract

As a novel approach we will combine trajectories or longitudinal studies of gene expression with information on annual influenza epidemics. Seasonality of gene expression in immune cells from blood could be a consequence of within-host seasonal immunity interacting with the seasonal pandemics of influenza (flu) in temperate regions and, thus, with potential valuable analogy transfer to the proposed seasonal development of covid-19.

Here we operationalized within-host immunity as genes with both a significant seasonal term and a significant flu term in the sine-cosine model. Information on gene expression was based on microarray using RNase buffered blood samples collected randomly from a population-based cohort of Norwegian middle-aged women in 2003-2006, The Norwegian Women and Cancer (NOWAC) study. The unique discovery (N=425) and replication (N=432) design were based on identical sampling and preprocessing. Data on proportion of sick leaves due to flu, and the flu intensities per week was obtained from the National Institute of Public Health, giving a semi-ecological analysis.

The discovery analysis found 2942 (48.1%) significant genes in a generalized seasonal model over four years. For 1051 within-host genes both the seasonal and the flu term were significant. These genes followed closely the flu intensities. The trajectories showed slightly more genes with a maximum in early winter than in late summer. Moving the flu intensity forward in time indicated a better fit 3-4 weeks before the observed influenza. In the replication analyses, 369 genes (35.1% of 1051) were significant. Exclusion of genes with unknown functions and with more than a season in difference reduced the number of genes in the discovery dataset to 305, illustrating the variability in the measurements and the problem in assessing weak biological relationships. Thus, we found for the first time a clear seasonality in gene expression with marked responses to the annual seasonal influenza in a unique discovery – replication design. Hypothetically, this could support the within-host seasonal immunity concept.

## Introduction

Regular recurrences of virus epidemics and sometimes pandemics like seasonal influenza, flu, have raised the question of within-host seasonal immunity as an evolutionary adaptation of the human immune systems to upcoming seasonal infections (1). Temperature, humidity, and population density have been described over many centuries as important factors for spread of the infections (2). The first wave of covid-19 in winter 2020 showed evidence of a seasonal disease (3, 4). The third wave in 2022 in temperate regions of Europe and USA indicate that covid-19 behave as a seasonal virus (5). The seasonality of covid-19 is clearly demonstrated in the Norwegian number of deaths for the period February 2020 till January 2022, Figure 1 (6). The two diseases share the common nature of being mRNA virus but with important differences like influenza virus with negative sense and coronavirus with positive sense, among others (7, 8). These are two of the major zoonotic respiratory viruses with capabilities of a pandemic spread. Diagnostic tests based on gene expression from white blood cells at time of serious clinical disease have shown similarities between the two diseases, but also unique patterns of dysregulated expression compared to healthy state (9, 10). The last mutation SARS-CoV-2 omicron (11, 12) is more infectious, but with less clinical severity of infections than the first mutations of covid-19. Thus, covid-19 could emerge as a parallel to the seasonal influenza virus N1H1. A recent genomic study showed that a splice variant in OAS1 genes could be protective for COVID-19 (13), going back 60.000 years. There are indications of covid epidemics 25 000 years ago in Asia (14). Seasonal influenza had repeated mutations over the last centuries (7, 15). The pandemic in 1919 named the Spanish flu caused millions of deaths due to a N1H1 virus. The high mortality among young men during the first world war has been explained by a lack of immunity to those born after the 1889 “Russian flu”. However, the 1889 “Russian flu” pandemic has also been described potentially as a coronavirus epidemic that could give ideas about the future handling of a seasonal covid-19 (16).

**Figure 1.**
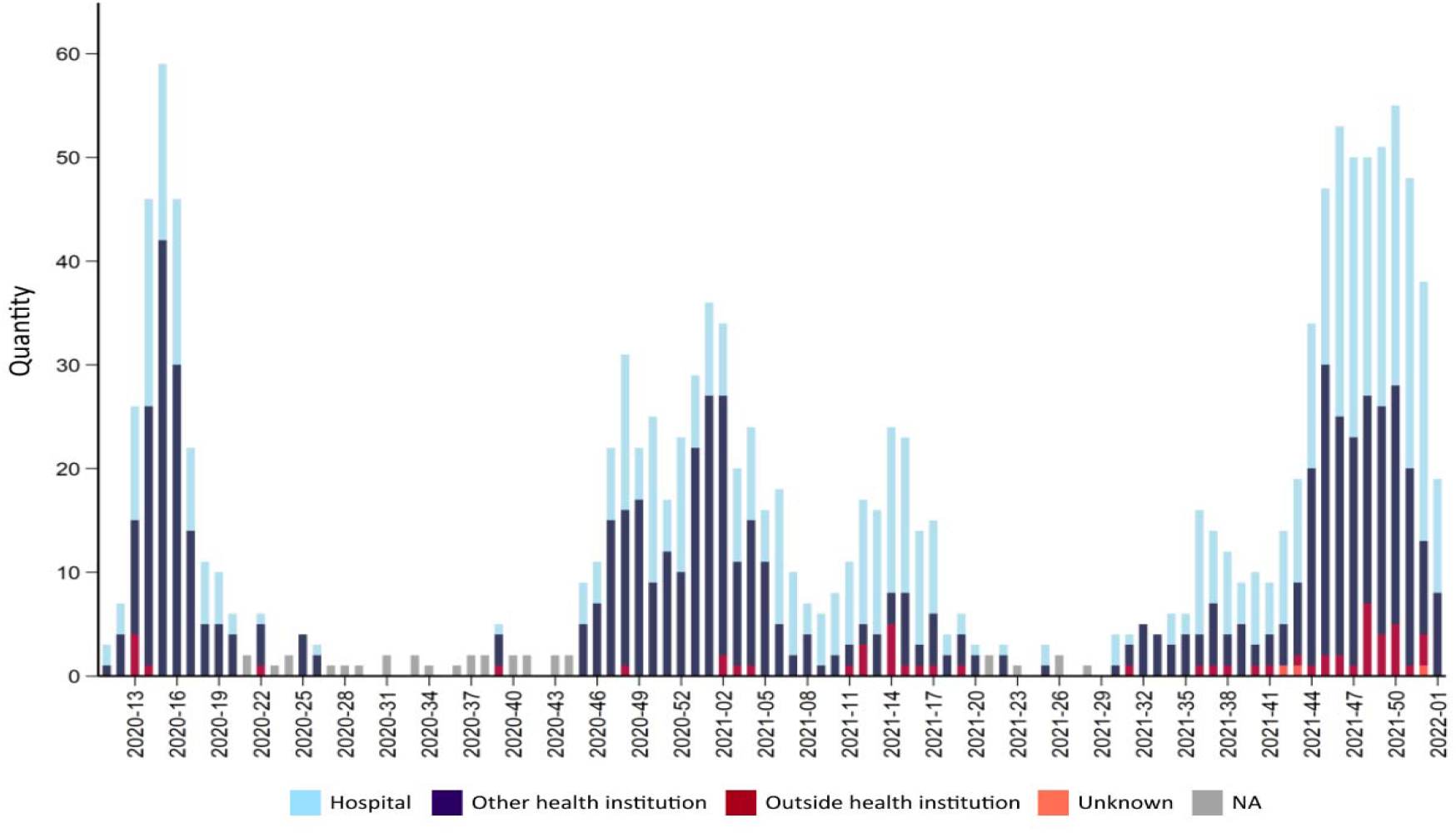
Number of deaths due to or with covid-19 for each week during the pandemic 2020-2022. Source FHI Weekly report 2022

This seasonality of virus infections during evolution could have changed the immune system in a seasonal direction. Several studies have shown seasonality of gene expression from immune cells (17, 18, 19) during the year. Some analyses included the effect of diagnosed virus infections but did not look at longitudinal effects of seasonal influenza. No general population studies have demonstrated repeated seasonality over years that also included information on seasonal influenza each year. The nature of this potential immunological mechanism is not defined nor any specific gene involvement. A huge number of analyses of other immune parameters have found both seasonal and daytime variations (20, 21).

Within-host seasonal immunity can be operationalized in sine-cosine models as genes have both significant seasonal effects and flu effects, in contrast to genes with either significant seasonal or significant flu effects (22). A within-host seasonal immunity would have to be a generalized effect covering most of a population. With the emerging knowledge of potentially two different seasonal virus pandemics, identification of gene expression profiles before and during the flu epidemic could be important for understanding a potentially seasonal covid-19 pandemic.

A search in MEDLINE combining seasonality, gene expression and influenza virus gave only 8 hits, see Box 1. Of the eight articles six were about vaccination, one clinical study and one animal experiment.

Here we will describe for the first time the changes or trajectories in gene expression before, at and after the annual seasonal flu epidemics. We will take advantage of the national surveillance system (23) of seasonal influenza epidemics in Norway that reports the proportion of sick leaves from influenza like illnesses (ILI) as proportion of all sick leaves. Figure 2A clearly shows the seasonal waves of influenza every winter, with the exception of 2004 when the epidemic came earlier. Gene expression analyses used RNase buffered whole blood samples from the adult Norwegian female population collected randomly in 2003-2006 as part of The Norwegian Women and Cancer study (NOWAC) postgenome biobank designed for gene expression analyses (24). A discovery - replication design will be implemented to improve reproducibility (25). The complicated explorative design focus on virology and epidemiology, postponing detailed pathways analyses.

**Figur 2A.**
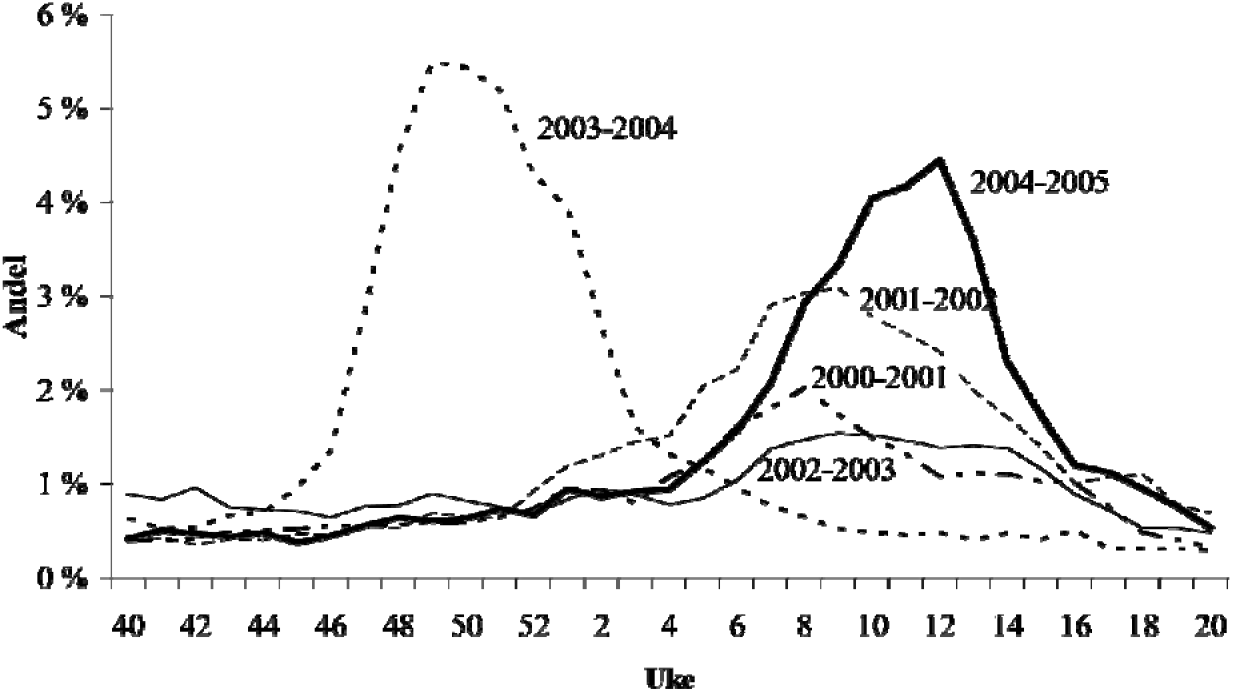
Percentage of the number of weekly consultations related to flu at general practitioners in Norway in the period 2000-2005, the Norwegian Surveillance System for Communicable Disease (MSIS) www.fhi.no/globalassets/dokumenterfiler/influensa/influensaovervaking-gml/sykdomsovervaking---influensasesongen-2004-2005-pdf.pdf

**Figure 2B.**
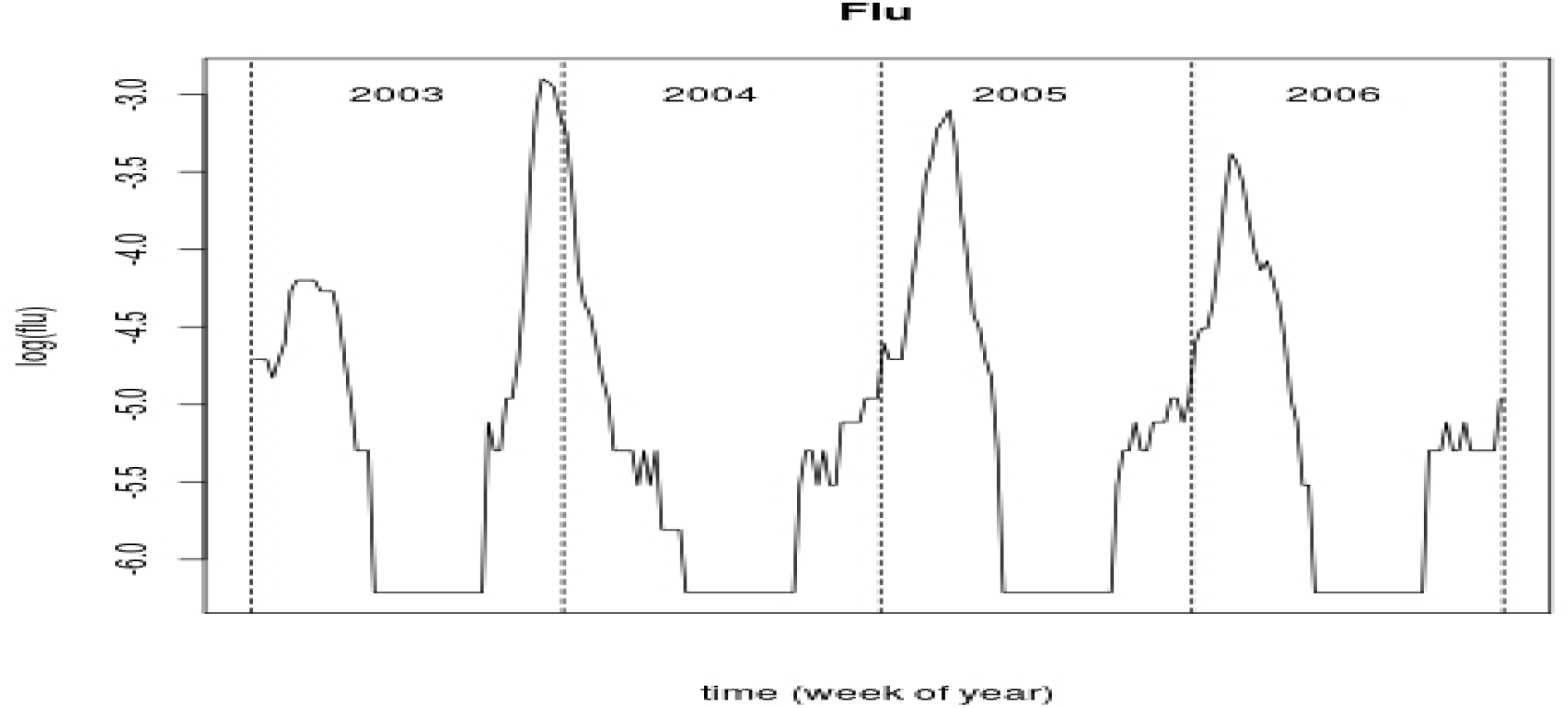
The figure shows the log of the weekly percentage of consultations at general practitioners in Norway, log(flu), related to flu in the study period 2003-2006. No reports were collected between week 20 and week 40, the middle of the year. Data delivered from the Norwegian Surveillance System for Communicable Disease (MSIS), numbers identical to figure 2A.

The aim here is to explore the trajectories of gene expression or seasonal changes in relation to the annual epidemics of seasonal influenza searching for genes expressing within-host seasonal immunity.

## Methods

### Study design and participants in NOWAC (Kvinner og kreft)

The NOWAC study is a prospective study with recruitment of 172 000 women randomly sampled from the National Population Register in Norway invited between 1991 and 2007 (26). These women received a letter of invitation and a questionnaire, and additional questionnaires were sent out with intervals of four to six years. All participating women were followed up through linkage to national cancer and death registries based on the unique national identification number assigned to all residents of Norway.

### Ethical issues

The NOWAC study was approved by the Norwegian Data Inspectorate and recommended by the Regional Ethical Committee of Northern Norway (REC North). The linkages of the NOWAC database to national registries such as the Cancer Registry of Norway and registries on death and emigration have also been approved. The women were informed about these linkages in the letter of invitation. Furthermore, the collection and storing of human biological material was approved by the REC North in accordance with the Norwegian Biobank Act. The linkages between Cancer Registry data and NOWAC study participants were performed at Statistics Norway, and the dataset was fully anonymized before it was made available to the authors. Information on breast cancer were used in this study to sample the random, matched controls. The Norwegian Data Protection Authority gave NOWAC exemption from the duty of confidentiality and permission to handle personal data (Datatilsynet, ref. 07/00030-2/cbr).

### Blood samples and data collection: The NOWAC Post-genome Cohort

In 2003-2006, NOWAC participants born 1943-1957 were invited to participate in a sub-cohort: the NOWAC Post-genome Cohort (24) with the Post-genome biobank. The main purpose of this cohort was to establish a biobank suitable for analyses of functional genomics, in particular transcriptomics. Random samples of NOWAC participants were drawn in weekly batches of 500, until 50,000 women participated in. This gave a random date for each blood donation. These women completed a two paged questionnaire. Whole blood samples were collected by a general practitioner or at a health care institution, using the PAXgene Blood RNA collection kit (Preanalytix/Qiagen, Hombrechtikon, Switzerland), and were transported to the Institute of Community Medicine at UiT in Tromsø by overnight post for biological analyses. The PAXgene Blood RNA collection kit contains a buffer that lyses the blood cells and preserves the mRNA profile of the sample, allowing for long-term frozen storage and optimizing the sensitivity of analyses (27). The sampling was stopped for 6 weeks in the Norwegian summer holidays mostly from end of June till the beginning of August, and around Christmas.

### Sampling strategy

These analyses were based on a split-sample strategy. Eligible Post-genome participants who were diagnosed with incident breast cancer were identified through linkage to the Cancer Registry of Norway. Each of these cases were assigned a matched control at random from the Post-genome cohort with the same birth year and same weekly batch of 500 invited women. Information on cases were not used in this study. The persons used as controls represented a part of the study population without any known cancer diseases. The discovery and replication populations were both controls from two case-control studies of breast cancer. The discovery population consisted of 425 women used as controls in an analysis of gene expression trajectories before time of diagnosis (28). The replication used 432 women that had been controls in a study of changes in gene expression trajectories after a diagnosis of breast cancer (29). The distribution of the two studies according to sampling year is shown in Table 1.

**Table 1.** Number of persons in each year for the Discovery and Replication datasets

|  | Study year, number of persons with blood sample |  |  |  |  |
| --- | --- | --- | --- | --- | --- |
| Dataset | 2003 | 2004 | 2005 | 2006 | Sum |
| Discovery | 94 | 107 | 148 | 76 | 425 |
| Replication | 96 | 28 | 112 | 196 | 432 |

### National influenza information

The information on seasonal influenza epidemics in Norway 2003-2006 was obtained by request from the MSIS system (Meldingssystem for smittsomme sykdomme) which is The National System for Notification of Infectious Diseases at The National Institute of Public Health (NIPH) (23). From autumn 1998 NIPH designated 201 general practices as sentinel reporting units based on geographical location, population size and previous reporting frequencies. These formed about 10% of the practices, but about 25% of the reported volume of influenza. The sentinels report weekly, from week 40 in autumn to week 20 in spring, the number of cases of ‘R80 Influenza’ from the International Classification of Primary Care (ICPC). Additionally, the number of consultations is reported. From 2004–2005 the number of patients on the patient list of general practitioners were used as denominators. The weekly recording period was from Friday to Thursday, after which the report card was completed and sent to NIPH. The national data provide a constant value for each week, here named *Flu intensity*. In the analyses weeks with no observations, the *Flu intensity* is set equal to 0.002 which is 2/3 of the smallest observed positive values. The data is smoothed with 15 days average.

### Laboratory procedures

All laboratory services were provided by the Genomics Core Facility, Norwegian University of Science and Technology, Trondheim, Norway. To control for technical variability such as different batches of reagents and kits, day-to-day variations, microarray production batches, and effects related to different laboratory operators, the original case-control pair was kept together throughout all extraction, amplification, and hybridization procedures. Total RNA extraction was performed using the PAXgene Blood RNA kit (Preanalytix/Qiagen, Hombrechtikon, Switzerland) according to the manufacturer’s instructions. RNA quality and purity were assessed using the NanoDrop ND 8000 spectrophotometer (ThermoFisher Scientific, Wilmington, DE, USA) and Agilent bioanalyzer (Agilent Technologies, Palo Alto, CA, USA). RNA amplification was performed on 96-wells plates using 300 ng of total RNA and the Illumina TotalPrep-96 RNA Amplification Kit (Ambio, Inc., Austin, TX, USA). The mRNA amplification procedure consisted of using a oligo(dT) primer for reverse transcription with a T7 promoter-specific ArrayScript reverse transcriptase, followed by a second-strand synthesis. *In vitro* transcription with T7 RNA polymerase using a biotin-NTP mix produced biotinylated cRNA copies of each mRNA in the sample.

The discovery samples were run on the Illumina Human Hu-6. The replication samples were run on the Illumina HumanHT-12 version 4 bead chip array (Illumina, San Diego, California, USA). The last one had half the beads of the first, see Appendix Figure A. Outliers were detected using the R-package nowaclean (30), based on visual examination of dendrograms, principal component analysis plots and density plots. Individuals that were considered borderline outliers were excluded if their laboratory quality measures were below given thresholds (RIN value <7, 260/280 ratio <2, 260/230 ratio <1.7, and 50 < RNA < 500). All laboratory work was done consecutively for all controls in 2012. The total number of genes were 25 212.

### Preprocessing of microarray data

The procedures for the preprocessing are given in (30). The discovery dataset consisted finally of 430 controls after exclusions of non-eligible and the replication set of 432 controls. The resulting datasets was then background corrected using negative control probes, log_2_ transformed using a variance stabilizing technique (31), and quantile normalized. We retained probes present in at least 90% of women. A probe was defined as present for an individual if its detection p□value was less than 0.01 for that individual. The set points for these two parameters were stricter than in previous analyses leaving fewer genes with small variations. If a gene was represented with more than one probe, the average expression of the probes was used as the expression value for the gene. The probes were translated to genes using the lumiHumanIDMapping database (32). The two largest principal components of the two data set were used for outlier control, see Appendix Figure B. We removed five observations from the original discovery data set that were outliers in the second principal component leaving 425 samples for the analyses. After preprocessing the discovery dataset had 6118 genes and the replication data set 6348 genes.

### Statistical models

In this Section we introduce several models where we can estimate a potential within-house seasonal and flu variation in the gene expression. We assume the log2 gene expression levels, X_g,c_, for person *c* and gene *g*. Here, *c* = 1,…,*M*, where M is the number of persons, and *g* = 1,…,*N*_*g*_, where *N*_*g*_ is the number of genes. For each person we have an observation date *t*_*c*_. **Model A - seasonality**.

We apply a standard sine-cosine model for modelling seasonality (22, 33, 34):

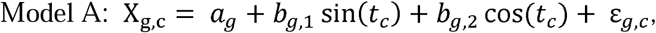

where ε_*g,c*_ ∼ *N*(0, *σ*_*g*_). We assume here that the date *t*_*c*_. is normalized to (− *π, π*) from July 1st one year to June 30th the year after.

The sine and cosine terms may express any shift in the sine function as may be seen from the equation: sin(*t* + *α*) = sin(*t*)cos(*α*) + cos(*t*)sin (*α*). This implies that we can reparametrize the model to:

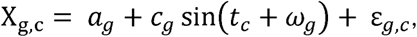

where 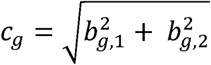 and *ω*_*g*_ = atan2(*b*_*g*, 2_, *b*_*g*, 2_) that also is normalized to (−*π, π*). We also introduce 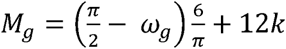 as a continuous variable for the month in the year where the integer *k* is selected such that 0 ≤ *M*_*g*_ ≤ 12. If the seasonal effect is largest in the beginning of January, 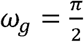 and *M*_*g*_ = 0. If the seasonal effect is largest in beginning of July, 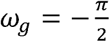 and *M*_*g*_ = 6. We may characterize *c*_*g*_ as log fold change since the maximum deviation from the average value of the seasonal variation is *c*_*g*_.

### Model B – seasonality and flu

We have weekly data for flu in the period 2003-2006. We denote the logarithm of these data as *f*(*t*). We obtain Model B by adding a flu term to model A:

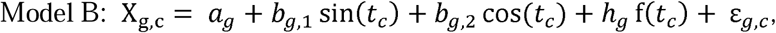

where *f*(*t*) is the logarithm of the ratio of consultations related to flu for general practitioners and *h*_*g*_ is the coefficient of the flu term. The function *f*(*t*) is smoothed and the minimum value is set equal to 0.002. The size and period of the seasonal term may be calculated from *b*_*g*,1_ and *b*_*g*,2_.

### Model C – seasonality and flu moved in time

It is of interest to find out whether the change in the gene expression is mainly coming earlier than, coinciding with or coming after the change in the observed flu intensity. We therefore introduce the model C:

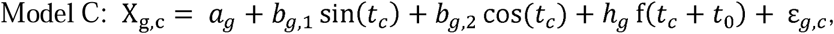

where *t*_0_ is a fixed constant moving the flu term forward or backward in time. We estimate the model for different values of *t*_0_ and observe the change in the number of significant genes as a function of *t*_0_.

### Hypothesis test – Is the flu effect due to a subgroup of individuals?

The effect of flu on gene expression may either be a large effect in a subgroup of individuals or a smaller effect on the entire population. We introduce a hypothesis test to separate between these two effects. This hypothesis test is based on the regression formula from Model B where the seasonal variation is omitted, i.e. *b*_*g*,1_ = *b*_*g*,2_ = 0, including only the flu term. If the correlation with the observed flu intensity is due to a subgroup of individuals, we expect that the residual term |ε_*g,c*_ | is large when the flu term | *h*_*g*_ f(*t*_*c*_)| is large and ε_*g,c*_ *h*_*g*_ > 0 for many genes for the same person, i.e. we expect more extreme gene expressions for this sub-group than the average gene expression. The null hypothesis is that the residual term ε_*g,c*_ is symmetric around zero and independent of the flu. Based on this, we introduce the statistics

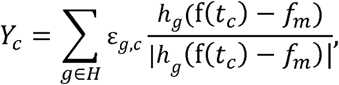

where the sum is over the set of genes *H* where the FDR-corrected p-value for a flu term in Model B is below 0.05 and | *h*_*g*_ | >0.1. The constant *f*_*m*_ is the average of *f*(*t*_*c*_) for all the individuals. If a subgroup of individuals contributes to a significant flu term, we expect a limited number of large values for *Y*_*c*_ when | *f*(*t*_*c*_)−*f*_*m*_| is large. Under the null hypothesis however, we expect *Y*_*c*_ to be symmetric around zero and close to a normal density. If the residual has higher variability when | *f*(*t*_*c*_)−*f*_*m*_| is large, then we would expect the same number of small negative values of *Y*_*c*_ as large values for *Y*_*c*_ when | *f*(*t*_*c*_)−*f*_*m*_| is large.

Let *V*_(*i*)_ denote the variable *V*_*c*_ = f(*t*_*c*_)−*f*_*m*_ where this variable is sorted after decreasing order of *Y*_*c*_. Note that all large values of | f(*t*_*c*_)−*f*_*m*_ | has f(*t*_*c*_)−*f*_*m*_ >0. Under the null hypothesis, *V*_*c*_ is independent of *Y*_*c*_, while if the covariation of the flu and gene expression is due to a sub-group of individuals, both *Y*_*c*_ and *V*_*c*_ will be large for these individuals. Based on the definitions above, we define the test statistic

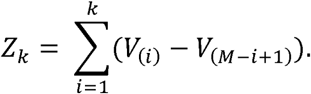

We observe that *Z*_*k*_ is the difference between the sum of f(*t*_*c*_)−*f*_*m*_ for the *k* largest values of *Y*_*c*_ and the *k* smallest values of *Y*_*c*_. *M* is the number of persons. Under the null hypothesis we expect *Z*_*k*_ to be close to zero since *V*_*c*_ and *Y*_*c*_ are independent, while under the alternative hypothesis we expect large values of *V*_*c*_ when *Y*_*c*_ is large implying large values of *Z*_*k*_. We therefore use *Z*_*k*_ as test statistic for the null hypothesis that the residual ε_*g,c*_ is symmetric around zero, while the alternative hypothesis is that there is a subgroup of individuals that have large values of ε_*g,c*_ when |*h*_*g*_ (f(*t*_*c*_)−*f*_*m*_)| is large. We compute the null distribution of *Z*_*k*_ by permuting the values of *V*_*c*_ and reject the null hypothesis if *Z*_*k*_ is larger than expected by chance.

## Results

We used the Discovery and Replication datasets in Table 1 for estimating the parameters in the models and testing the hypothesis described in Statistical Models.

### Model A – seasonality

The model with only a seasonal term, model A, was applied on the Discovery dataset. This gave 2942 significant genes of 6118 genes (FDR 5%). Of the 2942 significant genes, 416 had log fold change (*c*_*g*_) larger than 0.2. Figure 3 shows the seasonal variation of the 2942 genes. There were clear seasonal effects both in a summer and a winter season. Note that there were few observations during summer vacation and at the end of December.

**Figure 3.**
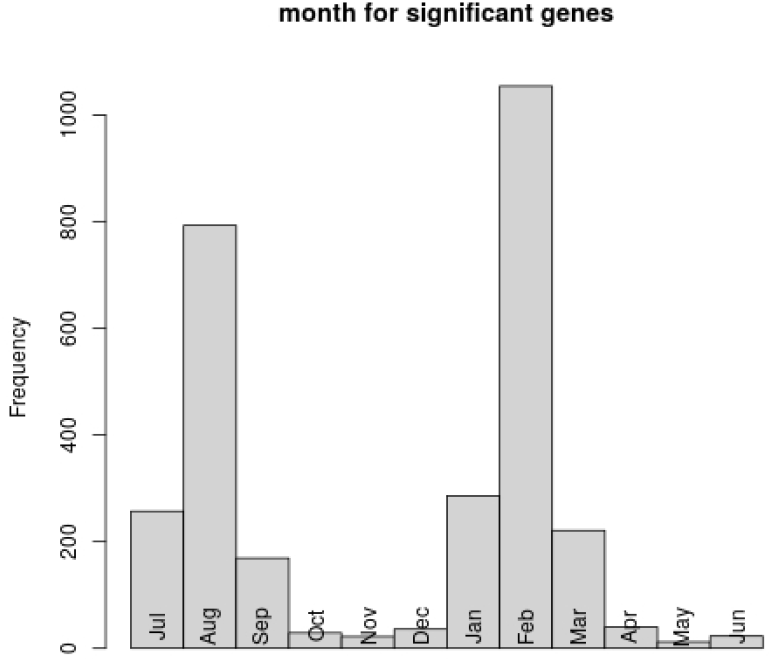
The distribution of the month M_g_ where the estimated gene expressions in the Discovery dataset are strongest for the 2942 genes with significant seasonal variation in Model A with seasonality.

### Model B – seasonality and flu

As shown in Figure 2B, the flu intensities had a marked seasonality with up to 5% of all sick leaves due to ILI. The maximum value of the flu varied between the years. In 2002-2003, the maximum was in February - March 2003. In 2003-2004 the epidemic came as early as the beginning of December 2003. This looked like two epidemics in the same year. Then the maximum in 2004-2005 came more than a year later in middle of March 2005. In 2005-2006 the maximum was during winter.

The result from Model A and Model B for the Discovery dataset are shown in a Venn diagram in Figure 4. The area denoted Season 1 show the number of genes in Model A where at least one of the seasonal parameters *b*_*g*,1_ and *b*_*g*,2_ are significant. Correspondingly, the area denoted Flu 1 counts the number of significant genes in Model B where the seasonal variation is omitted, i.e. *b*_*g*,1_ = *b*_*g*,2_ = 0 and where the flu parameter *h*_*g*_ is significant. The areas denoted Season 2 and Flu 2 show the number of genes in Model B where, respectively, at least one of the seasonal parameters *b*_*g*,1_ and *b*_*g*,2_ is significant and the flu parameter *h*_*g*_ is significant.

**Figure 4.**
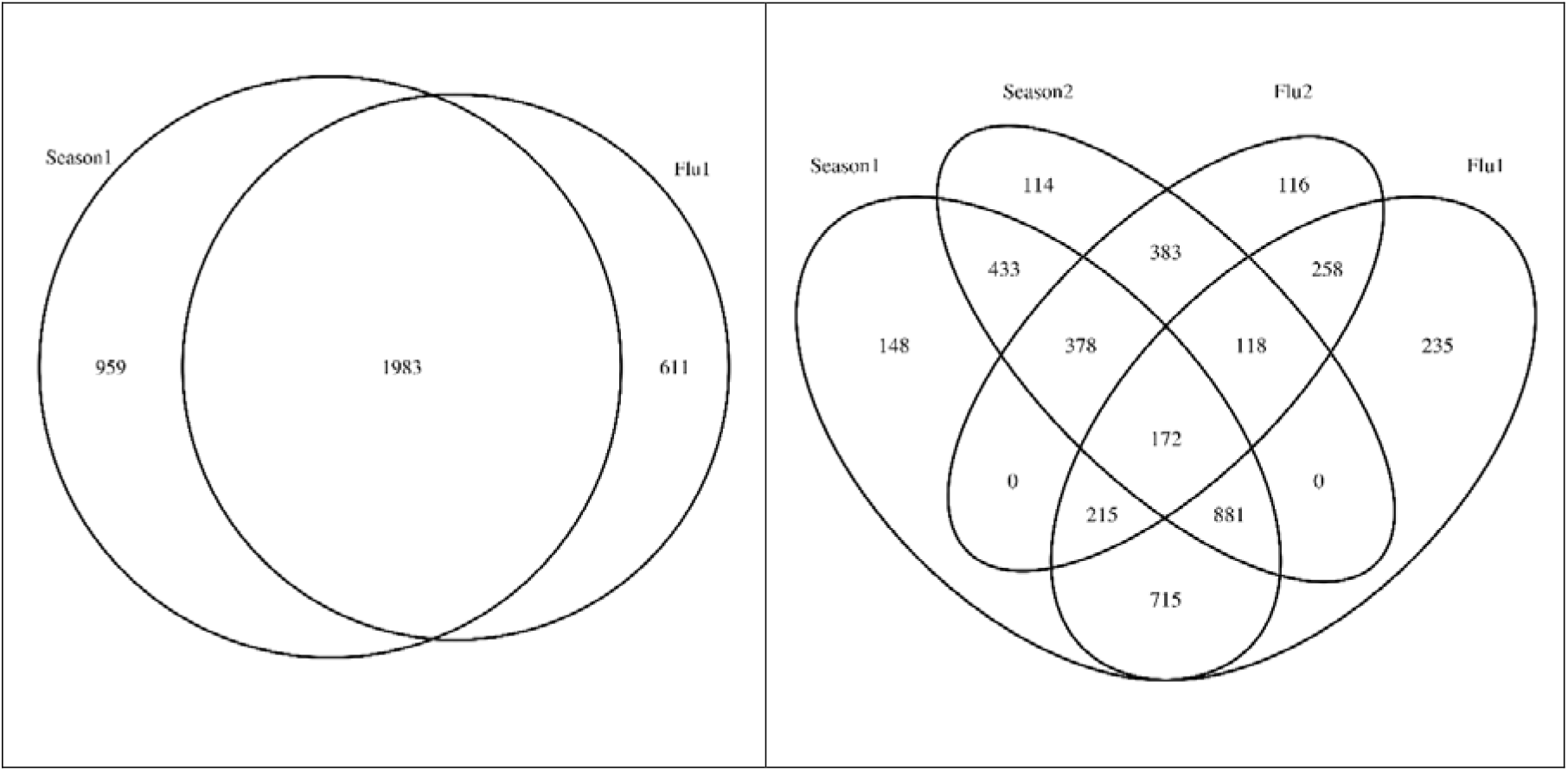
Venn diagram with the number of significant genes from the Discovery dataset when there is only a seasonal term (Season 1, Model A), only a flu term (Flu 1, Model B with b_g,1_ = b_g,2_ = 0) or a significant seasonal term (Seasonal 2) or a significant flu term (Flu 2) when the model has both terms (Model B).

The left panel in Figure 4 shows that mainly the same genes (1983 genes) have a significant seasonal term or a flu term when the model only includes one of the two terms. 959 genes only have a significant seasonal term, and 611 genes only have a significant flu term. Hence these terms mainly describe the same variability in the gene expressions. The seasonal term has the flexibility to set the maximum gene expression at any date during the year. For most genes the estimated season has its maximum or minimum value in February which is close to the average annual maximum value of the flu-observations. Hence, the *seasonal variation* for most genes may be explained by the flu variation. This indicates that the flu intensity may be a main driver in determining the seasonal variability of the gene expression. Table 2 shows that it is slightly more usual that the genes are up regulated with the flu (1488 genes) than down regulated (1106 genes) in Model B with b_g,1_ = b_g,2_ = 0.

**Table 2.** The sign of the significant flu coefficients h_g_ in Model B in the Discovery dataset. This model has seasonality (however this is turned off with b_g,1_ = b_g,2_ = 0) and a flu term.

| | $h_g < 0$ | $h_g > 0$ |
| --- | --- | --- |
| Number of significant genes | 1106 | 1488 |

The right panel in Figure 4 shows that 695 genes (114+433+148) in the discovery dataset have only a significant seasonal term, 609 only a significant flu term (116+258+235) while 1051 genes (378+383+172+118) have *both* a significant seasonal and a significant flu term when using Model B. Figure 5 describes the joint effect of the seasonal term and the flu term for the 1051 genes where both terms are significant in Model B. The upper panel compares the strength of the seasonal term and the flu term. There is a trend that genes with a strong flu trend also have a strong seasonal term. The sine function varies from -1 to 1, while the flu term varies from -6 to -3, see variation at vertical axis in the upper panel. Therefore, it is necessary for a seasonal coefficient to be 1.5 times as large in order to have the same effect. The ratios observed in Figure 5, upper panel, when comparing the values at the two axes indicate that *the seasonal variation is slightly dominating in the estimate for most genes*. Figure 5, lower panels shows that the genes that are reduced in the flu season, *h*_*g*_ < 0, have a positive seasonal term in the winter with a maximum value most frequently in February. Similarly, the genes that are increased in the flu season, *h*_*g*_ > 0, have a seasonal term with a maximum value that is most frequent in July and August.

**Figure 5.**
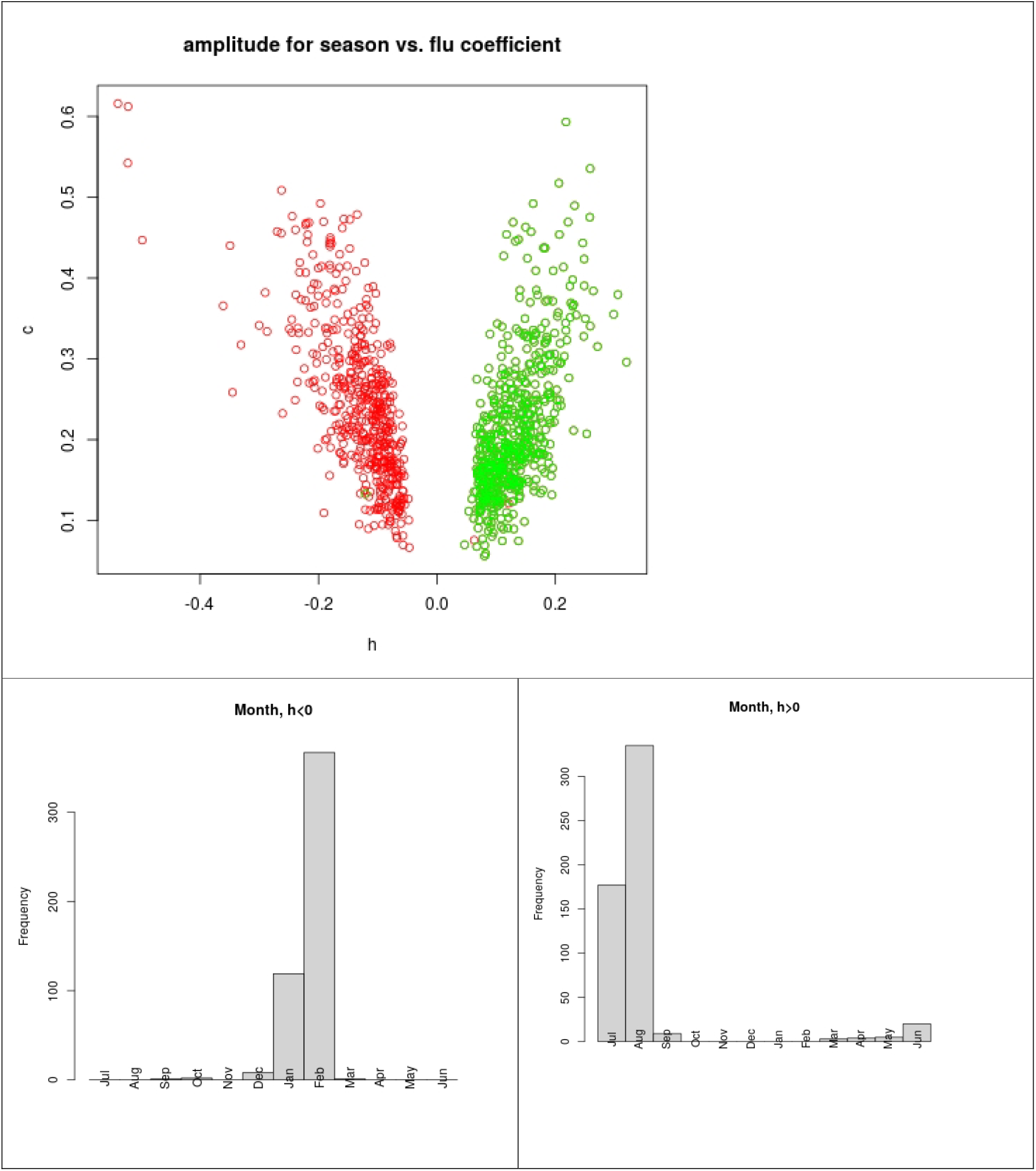
The upper panel show a cross plot of the flu coefficient h_g_ versus the amplitude c_g_ of the seasonal term in Model B for the 1051 genes where both these are significant in the Discovery dataset. Green circles indicate 3 ≤ M_g_ ≤9 and red circles indicate 0 ≤ M_g_ ≤ 3 and 9 ≤ M_g_ ≤ 12. Note that the season term is large when the flu term is small and the other way around. The lower panel shows the distribution of the month M_g_ with the maximum seasonal effect in Model B for the 1051 genes where both the seasonal term and the flu term are significant.

Figure 6 shows the estimated gene expression from Model A and Model B for three genes with the smallest *h*_*g*_ <0 and three genes with largest *h*_*g*_ >0. These six genes were selected from the 1051 genes with both a significant flu and seasonal effect in the four years period. For Model B with both a seasonal and flu term, the seasonal trend can be identified in the summer where the flu trend is constant. For the three genes with *h*_*g*_ >0, the seasonal trend has a maximum during the summer which is quite close to the top of seasonal trend in Model A. For the three genes with *h*_*g*_ <0, the seasonal trend has a minimum during the summer. In the rest of the year the variation of the flu changed over the four years. Notice that for two of the genes with *h*_*g*_ >0, the estimate is almost constant in Model A, and far from constant in Model B when a flu term was added and where the seasonal term also is large when there is a flu term.

**Figure 6.**
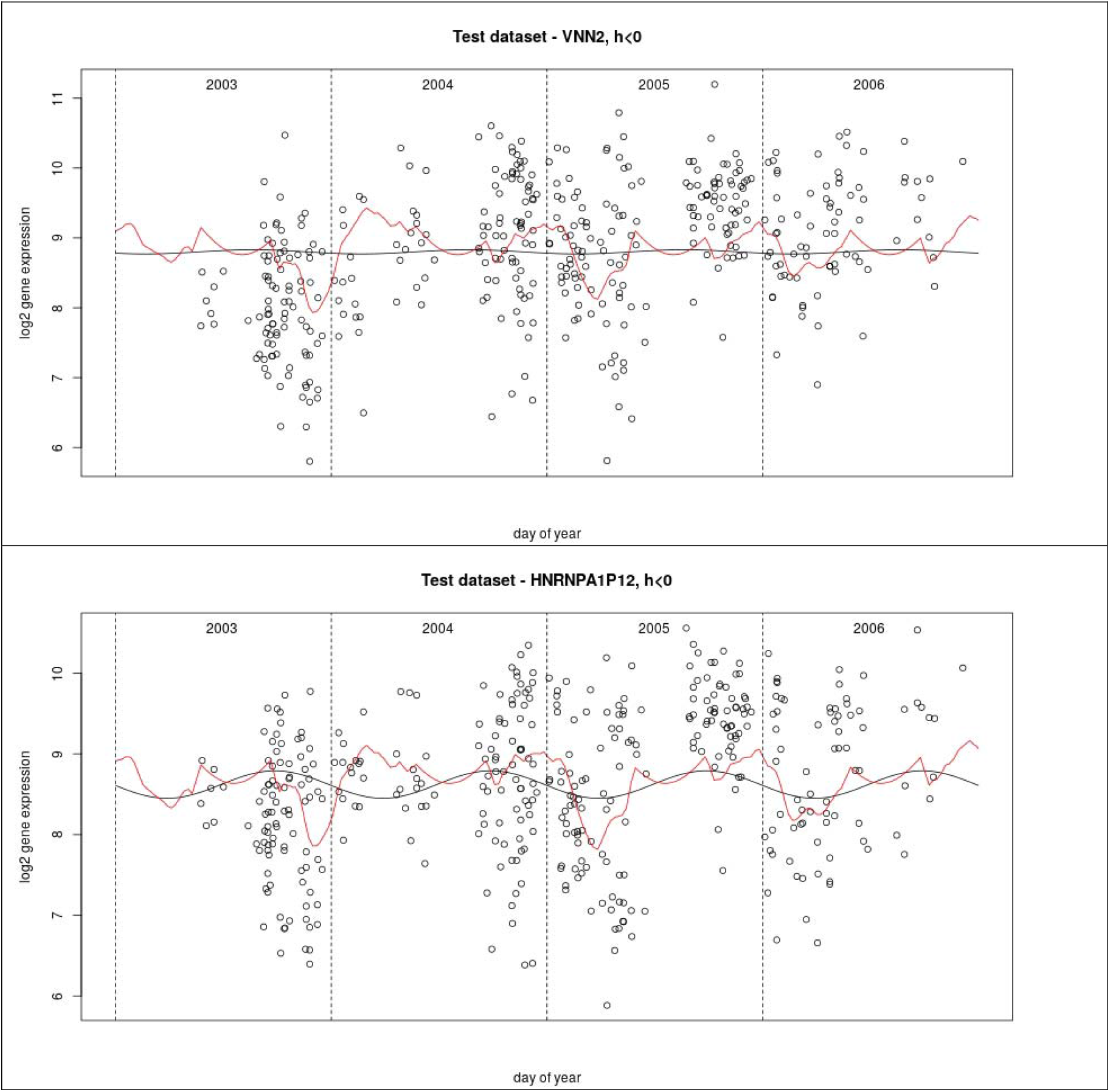
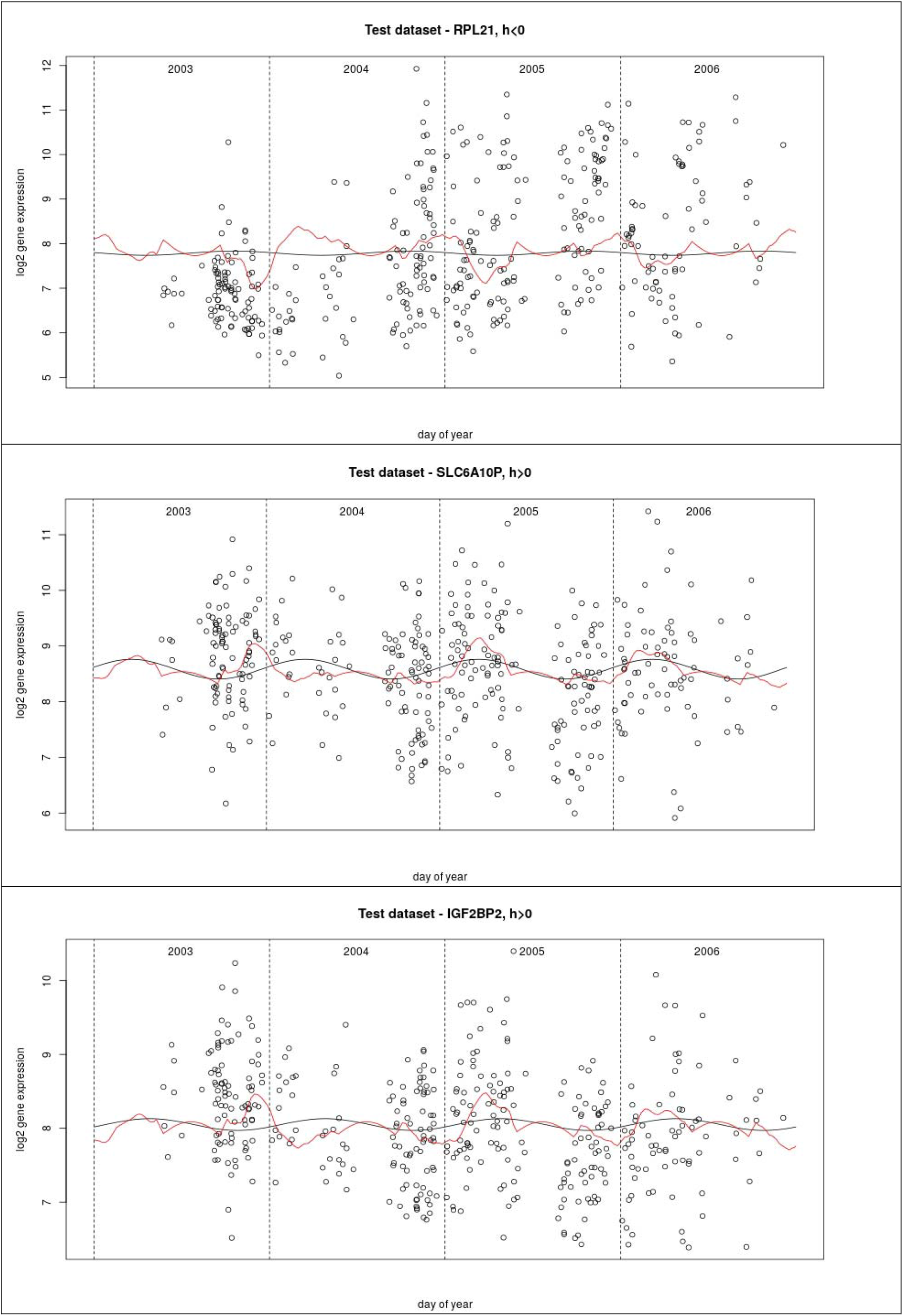
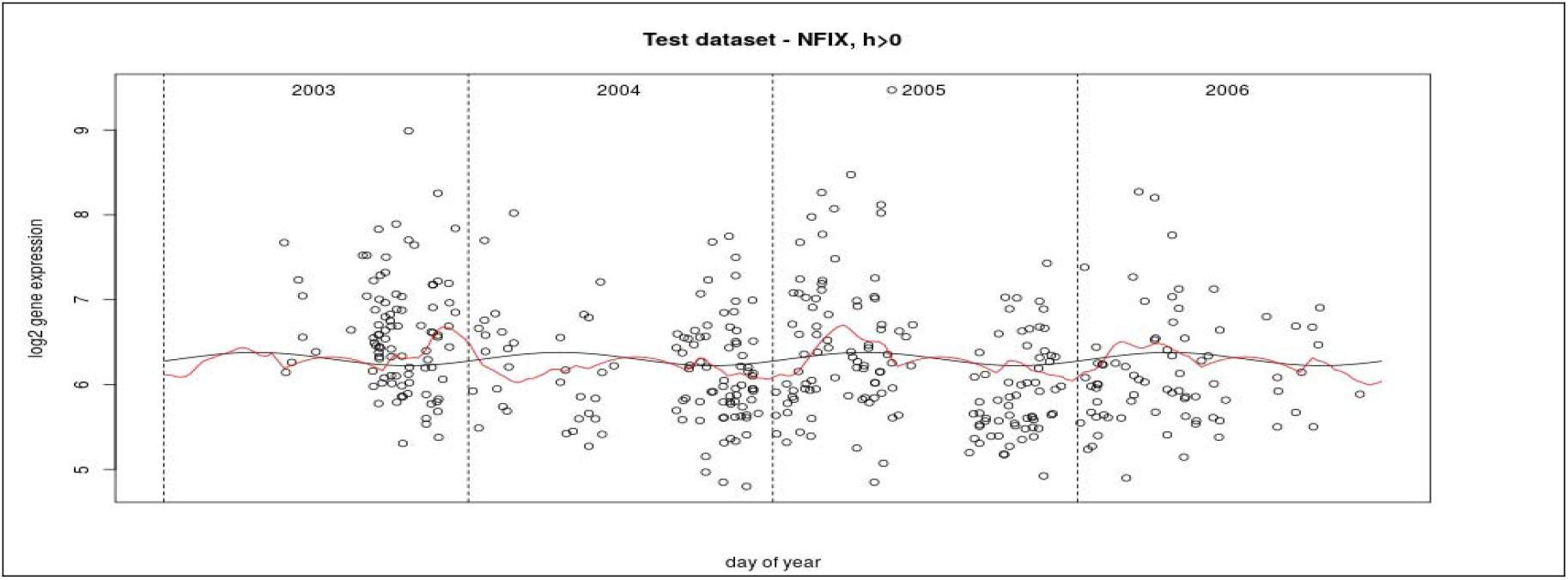
Estimated gene expression for three genes with smallest *h*_*g*_ < 0 and three genes with largest *h*_*g*_ > 0. *h*_*g*_ is the coefficient to the log flu intensity. The six genes are selected from the 1051 genes with both a significant flu and seasonal effect for the period 2003-2006, for the Discovery dataset. The estimate in Model A (season) is shown in black, the estimate in Model B (season and flu) is shown in red, and all observations are shown as circles.

The relationships between the flu intensities over the four years and the gene expression are shown in Figure 7. The smoothed log flu trend is shown together with the estimated gene expression from model B for the gene with largest and the gene with smallest *h*_*g*_ value, the coefficient for the flu term. The log fold change differences were for both in the order of 8.0 to 9.5, or a fold change of around 3.0. The changes in the flu coefficient closely resembled the estimate of the flu intensities either upwards or downwards. Notice the opposite sign of the season trend for the two genes. Since the seasonal terms most frequently has a maximum in January-February or July-August (with minimum in January-February), the seasonal term was after the top of the flu in 2003-2004 and before the top in 2004-2005 and 2005-2006.

**Figure 7.**
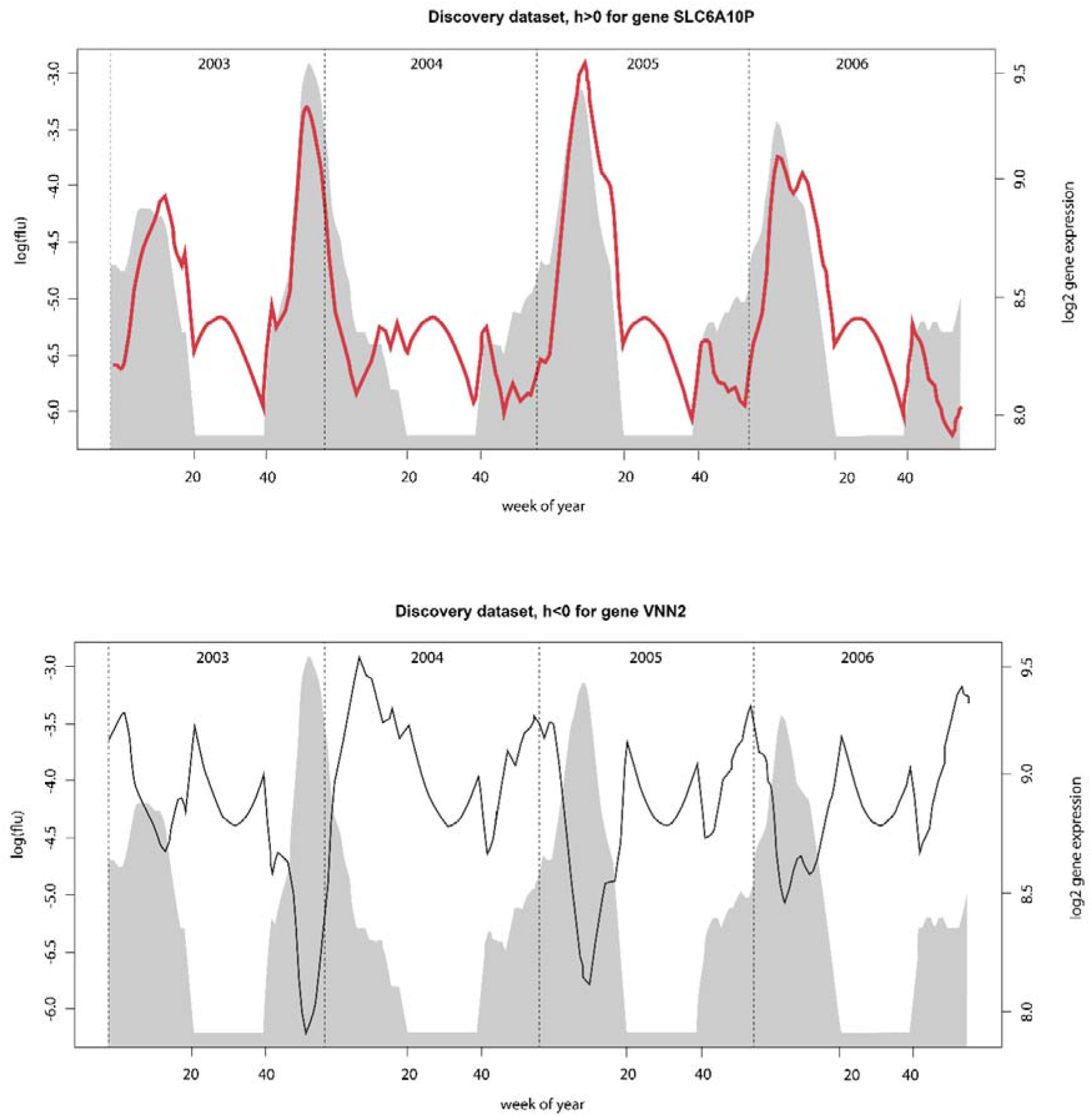
Gene expression trajectories for gene SLCA6A10P (largest *h*_*g*_ >0, upper panel, red line) and gene VNN2 (largest *h*_*g*_ <0, lower panel, black line) in relation to the estimated smoothed log flu intensities (grey shaded) 2003-2006

### Model C – seasonality and flu moved in time

Model C with both a season and a flu term were used for moving the flu intensities trend with *t*_0_ days, −60 < *t*_0_ < 60, to find out whether the change in the gene expression was mainly before, at the same time or after the changes in the flu intensity. The top panel in Figure 8 shows that the number genes with significant flu term or significant flu and at least one seasonal term were largest when the flu term was moved around three weeks forward. The number of genes with significant seasonal terms and no flu term was lowest when the flu term was 5 days later than the actual data. In total, this indicates that the main change in the genes occurred before the increase in the flu sick leaves. This was confirmed in the lower right panel showing that all the genes most downregulated obtained their minimum negative value around – 20 days. The movement of the upregulated genes were more complex; some genes obtained their optimum in the period (−25, -20) days, other genes obtained the optimum around 20 days and a final group obtained their optimum later in the spring. Table 3 is divided into sections marked with boldface for when the genes obtained the most extreme values. All the listed genes have a significant flu term, but in most cases not in the entire period, (−60,60) days.

**Figure 8.**
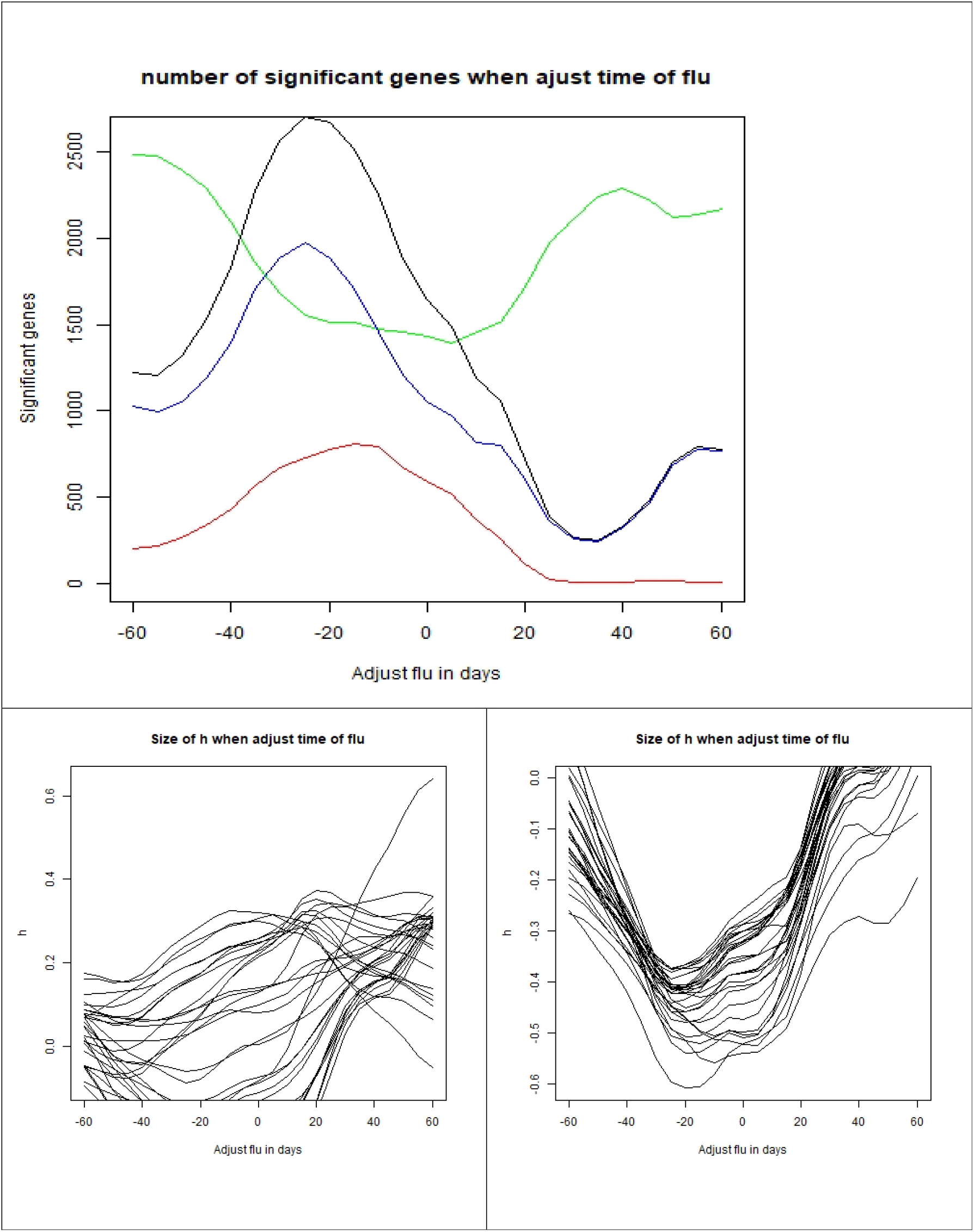
The effect of moving the flu trend forwards or backwards +/- 60 days in time, Discovery dataset, Model C. The top panel shows the number of genes with significant season/flu terms: flu term (black), both season and flu term (blue), flu term and not season term (red) and season term and not flu term (green). The lower left panel shows the size of coefficient for the flu term *h*_*g*_ when moving the flu term +/-60 days for the 30 genes with largest positive *h*_*g*_ values in the period. Lower right is similar for *h*_*g*_ < 0.

**Table 3.** The table shows how the coefficient of the flu term *h*_*g*_ changes when the flu intensity is moved +/-60 days in the Discovery dataset when using Model C with seasonality and a flu term. We show the estimated *h*_*g*_ -value and the corresponding FDR q-value when the flu term is moved -45 days (i.e. earlier), -15 days, 0 days, 15 days and 45 days. Period with smallest FDR q-value is written in boldface. The table is divided into sections. First there are three sections with *h*_*g*_ > 0 and then two sections with *h*_*g*_ < 0. Within each section, the genes are sorted after size of *h*_*g*_ where the most extreme value is obtained.

|  | -45 |  | -15 |  | 0 |  | 15 |  | 45 |  |
| --- | --- | --- | --- | --- | --- | --- | --- | --- | --- | --- |
| | $h_g$ | FDR q-value | $h_g$ | FDR q-value | $h_g$ | FDR q-value | $h_g$ | FDR q-value | $h_g$ | FDR q-value |
| SLC6A10P | 0.16 | 9.79E-02 | **0.29** | **1.02E-03** | 0.32 | 3.09E-03 | 0.29 | 2.81E-02 | 0.12 | 4.86E-01 |
| FCGR3A | -0.01 | 9.53E-01 | 0.12 | 1.40E-01 | 0.23 | 1.20E-02 | **0.35** | **1.22E-03** | 0.30 | 9.12E-03 |
| ACTRT1 | 0.06 | 4.80E-01 | 0.17 | 1.09E-02 | 0.26 | 1.93E-03 | **0.34** | **6.03E-04** | 0.28 | 8.70E-03 |
| DNA2 | 0.07 | 3.12E-01 | 0.18 | 5.38E-03 | 0.25 | 1.39E-03 | **0.32** | **5.16E-04** | 0.23 | 1.93E-02 |
| LOC653635 | 0.05 | 5.38E-01 | 0.17 | 2.03E-02 | 0.23 | 9.36E-03 | **0.32** | **2.30E-03** | 0.17 | 1.63E-01 |
| ADRA2C | -0.08 | 3.79E-01 | 0.07 | 3.68E-01 | 0.22 | 1.81E-02 | **0.31** | **4.23E-03** | 0.31 | 8.70E-03 |
| LOC399744 | 0.13 | 4.87E-02 | 0.26 | 2.16E-05 | 0.31 | 4.87E-05 | **0.30** | **8.28E-04** | 0.06 | 6.64E-01 |
| DSCAM | 0.10 | 1.01E-01 | 0.20 | 4.94E-04 | 0.25 | 5.05E-04 | **0.30** | **5.44E-04** | 0.16 | 8.48E-02 |
| ARTN | -0.07 | 4.18E-01 | 0.08 | 2.97E-01 | 0.21 | 1.33E-02 | **0.29** | **4.47E-03** | 0.27 | 1.35E-02 |
| LOC729021 | 0.13 | 1.52E-02 | 0.21 | 9.77E-05 | 0.26 | 7.20E-05 | **0.29** | **2.71E-04** | 0.18 | 3.15E-02 |
| IGF2BP2 | 0.15 | 2.97E-02 | 0.25 | 2.65E-04 | 0.30 | 3.12E-04 | **0.27** | **6.51E-03** | 0.16 | 1.42E-01 |
| RNF213 | -0.04 | 7.34E-01 | 0.05 | 6.46E-01 | 0.13 | 3.09E-01 | 0.27 | 5.44E-02 | **0.36** | **1.29E-02** |
| YY1 | 0.07 | 3.23E-01 | 0.10 | 1.09E-01 | 0.14 | 5.83E-02 | 0.17 | 5.51E-02 | **0.26** | **3.70E-03** |
| LRP10 | 0.07 | 2.88E-01 | 0.10 | 9.21E-02 | 0.13 | 5.24E-02 | 0.19 | 1.93E-02 | **0.25** | **3.16E-03** |
| RPL21 | -0.37 | 5.67E-03 | **-0.61** | **2.72E-06** | -0.52 | 1.11E-03 | -0.47 | 1.52E-02 | -0.11 | 6.83E-01 |
| FTHL8 | -0.20 | 2.73E-01 | **-0.54** | **9.63E-04** | -0.51 | 1.22E-02 | -0.39 | 1.19E-01 | 0.01 | 9.76E-01 |
| VNN2 | -0.33 | 2.79E-04 | **-0.52** | **1.23E-08** | -0.54 | 5.85E-07 | -0.49 | 1.85E-04 | -0.29 | 3.93E-02 |
| FKSG30 | -0.29 | 2.96E-02 | **-0.52** | **4.13E-05** | -0.50 | 1.22E-03 | -0.41 | 2.88E-02 | -0.02 | 9.54E-01 |
| RPS28 | -0.20 | 2.59E-01 | **-0.51** | **9.99E-04** | -0.47 | 1.37E-02 | -0.34 | 1.45E-01 | 0.08 | 8.13E-01 |
| LOC653658 | -0.23 | 1.62E-01 | **-0.48** | **1.68E-03** | -0.41 | 3.09E-02 | -0.33 | 1.54E-01 | 0.07 | 8.42E-01 |
| LOC390354 | -0.22 | 8.78E-02 | **-0.47** | **9.53E-05** | -0.44 | 3.35E-03 | -0.36 | 4.72E-02 | -0.04 | 8.96E-01 |
| LOC642817 | -0.31 | 2.77E-04 | **-0.47** | **1.60E-08** | -0.50 | 7.51E-07 | -0.40 | 8.87E-04 | -0.11 | 5.01E-01 |
| LOC728481 | -0.27 | 2.02E-02 | **-0.46** | **3.32E-05** | -0.38 | 5.85E-03 | -0.30 | 7.48E-02 | 0.08 | 7.48E-01 |
| FTHL12 | -0.21 | 1.50E-01 | **-0.46** | **4.62E-04** | -0.40 | 1.43E-02 | -0.29 | 1.40E-01 | 0.05 | 8.72E-01 |
| LOC389787 | -0.13 | 4.28E-01 | **-0.45** | **1.55E-03** | -0.38 | 3.33E-02 | -0.31 | 1.60E-01 | 0.02 | 9.64E-01 |
| RPL37A | -0.30 | 1.02E-02 | **-0.43** | **1.03E-04** | -0.32 | 2.22E-02 | -0.22 | 1.99E-01 | 0.15 | 4.70E-01 |
| HNRPA2B1 | -0.27 | 2.13E-02 | **-0.42** | **1.39E-04** | -0.41 | 3.13E-03 | -0.35 | 3.66E-02 | 0.00 | 9.87E-01 |
| CD48 | -0.17 | 2.50E-01 | **-0.42** | **1.22E-03** | -0.38 | 1.80E-02 | -0.34 | 8.51E-02 | 0.04 | 9.12E-01 |
| WASPIP | -0.24 | 4.73E-02 | **-0.42** | **3.01E-04** | -0.35 | 1.67E-02 | -0.32 | 6.96E-02 | -0.01 | 9.74E-01 |
| LOC645138 | -0.21 | 1.20E-01 | **-0.42** | **7.32E-04** | -0.33 | 3.58E-02 | -0.22 | 2.46E-01 | 0.16 | 4.88E-01 |
| AMY1C | -0.22 | 1.19E-01 | **-0.41** | **1.35E-03** | -0.36 | 2.71E-02 | -0.28 | 1.49E-01 | 0.06 | 8.53E-01 |
| RPS3A | -0.14 | 3.66E-01 | **-0.41** | **2.27E-03** | -0.35 | 3.82E-02 | -0.32 | 1.14E-01 | 0.06 | 8.42E-01 |
| UQCRHL | -0.23 | 6.25E-02 | **-0.41** | **4.98E-04** | -0.29 | 5.74E-02 | -0.22 | 2.17E-01 | 0.09 | 7.27E-01 |
| TM9SF2 | -0.26 | 3.18E-02 | **-0.41** | **3.64E-04** | -0.30 | 3.87E-02 | -0.24 | 1.78E-01 | 0.03 | 9.07E-01 |
| CDC42SE2 | -0.24 | 6.54E-02 | **-0.41** | **1.02E-03** | -0.28 | 8.08E-02 | -0.22 | 2.68E-01 | 0.07 | 7.91E-01 |
| FAM10A4 | -0.27 | 6.77E-03 | **-0.41** | **1.50E-05** | -0.32 | 6.96E-03 | -0.22 | 1.36E-01 | 0.07 | 7.45E-01 |
| LOC643310 | -0.26 | 7.61E-03 | **-0.41** | **1.78E-05** | -0.32 | 6.66E-03 | -0.22 | 1.26E-01 | 0.05 | 8.15E-01 |
| ATF4 | -0.22 | 9.63E-02 | **-0.38** | **2.20E-03** | -0.31 | 5.06E-02 | -0.24 | 2.21E-01 | 0.12 | 6.46E-01 |
| CLEC2D | -0.25 | 4.59E-02 | **-0.38** | **9.43E-04** | -0.30 | 4.18E-02 | -0.29 | 9.95E-02 | 0.10 | 6.95E-01 |
| LDHB | -0.25 | 1.65E-02 | **-0.37** | **1.76E-04** | -0.32 | 9.92E-03 | -0.24 | 1.07E-01 | 0.09 | 6.62E-01 |
| ZNF217 | -0.27 | 3.01E-03 | **-0.37** | **1.75E-05** | -0.29 | 8.39E-03 | -0.25 | 6.19E-02 | 0.01 | 9.74E-01 |
| PGAM4 | -0.25 | 1.50E-02 | **-0.37** | **1.33E-04** | -0.30 | 1.37E-02 | -0.24 | 1.11E-01 | 0.02 | 9.39E-01 |
| ARPC5 | -0.21 | 1.11E-01 | **-0.37** | **2.25E-03** | -0.30 | 4.88E-02 | -0.23 | 2.13E-01 | 0.12 | 6.10E-01 |
| SFRS10 | -0.25 | 1.50E-02 | **-0.37** | **1.60E-04** | -0.26 | 3.70E-02 | -0.19 | 1.98E-01 | 0.10 | 6.27E-01 |
| LOC648210 | -0.31 | 3.47E-04 | -0.47 | 2.72E-08 | -0.52 | 5.65E-07 | **-0.44** | **3.96E-04** | -0.15 | 3.33E-01 |

### Hypothesis test – Is the flu effect due to a subgroup of individuals?

We tested the hypothesis whether the covariation of the gene expressions and the flu intensity was due to a subgroup of individuals. In the null hypothesis, we assumed that the residuals ε_*g,c*,_ were symmetric around zero with the alternative hypothesis that we obtain large values of the residual when ε_*g,c*_*h*_*g*_ >0 is large. The results are shown in Table 4. We were not able to reject the hypothesis that the residuals are symmetric. Hence, we may not conclude that the observed correlation between the gene expression and the observed flu intensity was due to a subgroup of individuals. In the table, *k* represents the potential number of individuals with the most extreme residuals that potentially could dominate the covariation between the gene expressions and the flu intensity observations. The smaller *k* is, the more extreme must the residuals be to influence the covariation with the flu intensity for the entire sample.

**Table 4.** The probability for rejecting the hypothesis (p-value) that the residuals are symmetric based on 10,000 simulations. *k* represents the number of persons with most extreme residual values that potentially could dominate the covariation between the gene expressions and the flu intensity observations.

| $k$ | 10 | 20 | 30 | 40 |
| --- | --- | --- | --- | --- |
| $P(Z_k > \text{Observed value})$ | 0.20 | 0.59 | 0.58 | 0.42 |

### Replication analysis

To reduce noise in the data before interpretations of the genes, a replication analysis was performed for the 1051 within-host genes with both a significant seasonal and flu term. 87 genes from the Discovery dataset were not found in the Replication dataset. Of the remaining 964 genes 658 had either a significant season or flu term. In all, 369 genes had both a significant season and flu term. The upper panel in Figure 9 shows a cross-plot of the flu coefficient *h*_*g*_ against the amplitude *c*_*g*_ of the seasonal term in the Model B for the 369 genes that were significant for both in the replication dataset. There was a clear seasonality as shown in lower panel Figure 9, depending on the values of *h*_*g*_. When *h*_*g*_ <0 a January – March peak was found in contrast to genes with *h*_*g*_ >0 that had a peak in July – September.

**Figure 9.**
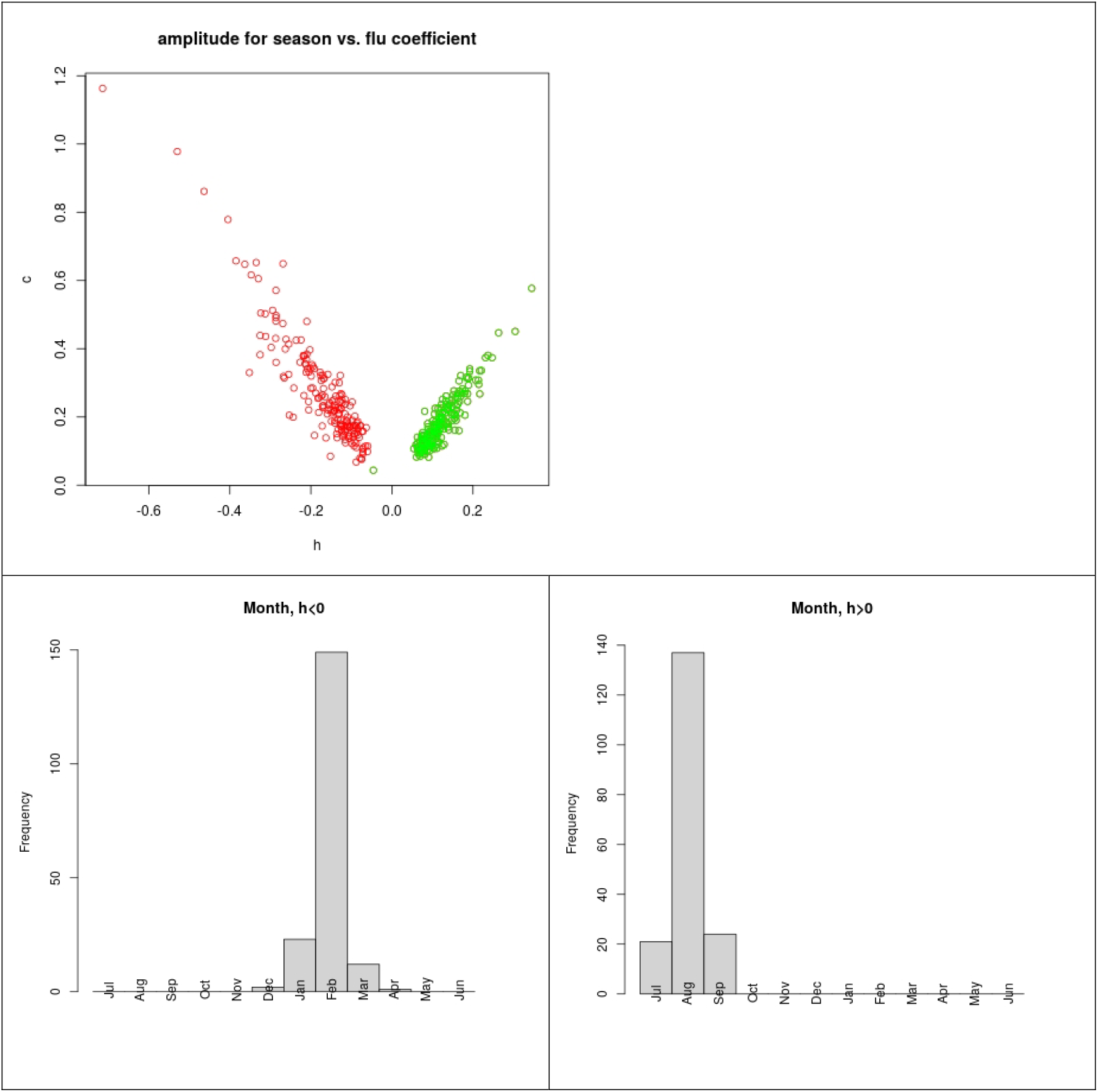
Upper panel shows a cross plot of the flu coefficient h_g_ versus the amplitude c_g_ of the seasonal term in Model B for the 369 genes where both these are significant in the Replication dataset. Green circles indicate 3 ≤ M_g_ ≤ 9 and red circles indicate 0 ≤ M_b_ ≤ 3 and 9 ≤ M_g_ ≤ 12. Note that the season term is large when the flu term is small and the other way around. Lower panel shows the distribution of the month M_g_ with the maximum seasonal effect in Model B for the 369 genes where both the seasonal term and the flu term are significant in the Replication data set.

**Figure 10.**
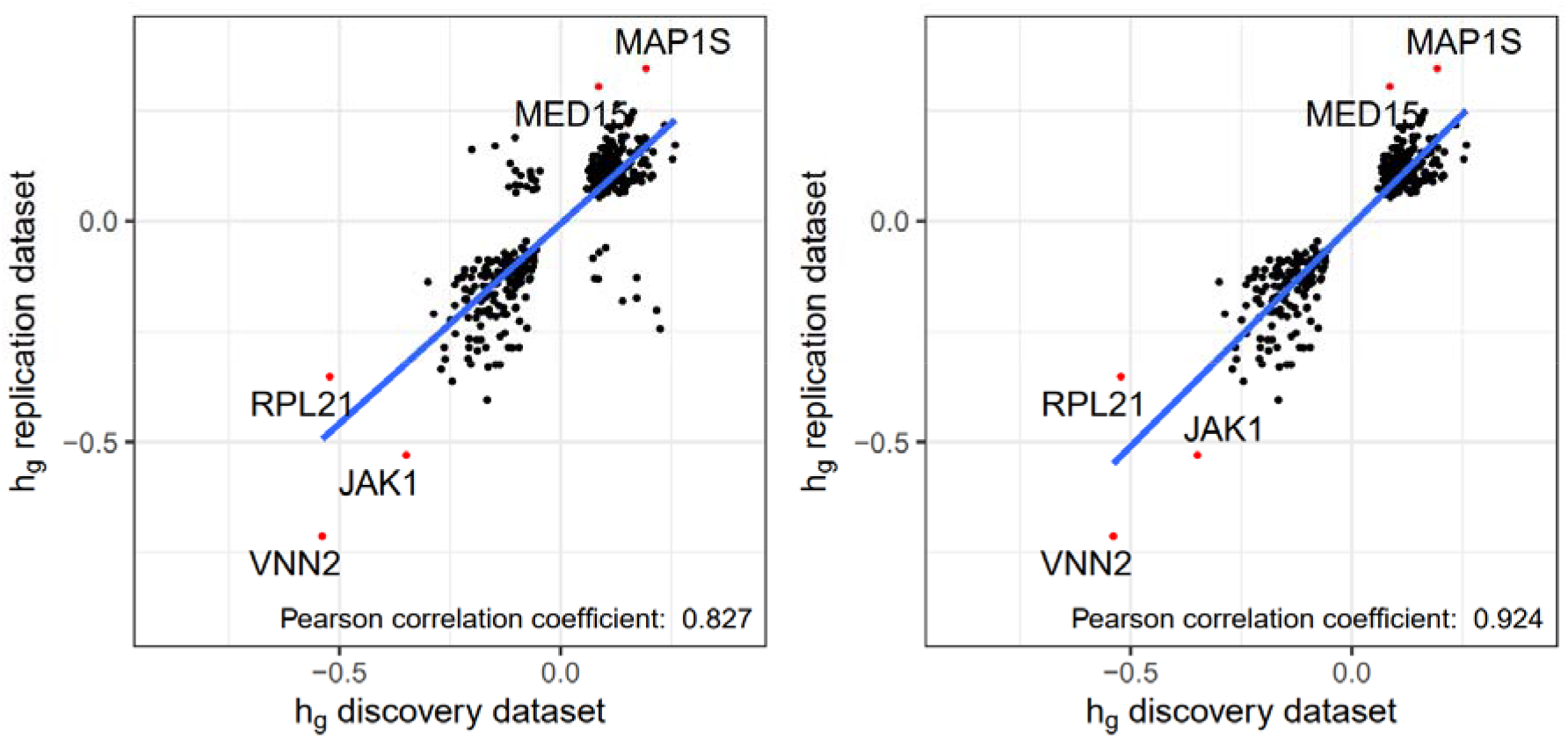
Correlation of values between the Discovery and Replication dataset. The red dots are the three most significant genes for <0 and the two most significant genes for >0. See Table 5A and Table 5B.

In the following, all genes with unknown function (LOC) in the Discovery dataset (N= 42) were excluded. We performed a correlation analysis of the months for the season between the Discovery dataset and the Replication dataset. When excluding 22 genes with more than one seasons difference in estimated month of maximum (FDR q-value <0.05), the Pearsons correlation coefficient changed from 0.83 to 0.92. This left 305 common genes, or 1.2% of all (305/ 25 212). This increased the Pearsons correlation coefficient for *h*_*g*_ from 0.71 to 0.98. Among the 305 genes, negative *h*_*g*_ values were found in 148 genes (Appendix Table 1) while 157 genes (Appendix Table 2) had positive *h*_*g*_ values. These genes represented two groups with distinct different seasonal trajectories of gene expression through the year. The flu varied from December to April. Those with positive *h*_*g*_ value had their maximum in the beginning of the year mostly overlapped by the flu. Those with a negative flu coefficient had maximum in the autumn.

### The function of the significant genes

A detailed in-depth investigation of the significant single genes and their functions was not among the aims of this work, but we include Tables 5A and 5B that summarize the functions of the top 20 genes for positive and negative flu coefficients, respectively. Additionally, we performed a pathway-level assessment of all significant genes associated with negative and positive *h*_*g*_ values using the Reactome database (data not shown). Generally, the number of significant pathways (p < 0.05) was higher for the *h*_*g*_ negative group of genes (263 against 49). Potentially, this can be explained by a more converged state of the immune system prepared for meeting oncoming infection.

**Table 5 A.** Gene names, function and immunological function of the 20 genes with highest positive value of *h*_*g*_.

| Gene | Function | Immunologic function |
| --- | --- | --- |
| ABCA7 | Plays a role in lipid homeostasis in cells of the immune system and in T-cell functions. Involved in Alzheimer | Participate in microglia immune response. |
| GNB2 | Encodes protein involved in transmembrane signaling. |  |
| SBNO2 | Involved in the transcriptional corepression of NF-kappaB in macrophages. Plays a role as a regulator in the proinflammatory cascade. Bone homeostasis | Involved in inflammatory responses |
| PA2G4 | Involved in growth regulation, ribosomal assembly. Transcriptional co-repressor of androgen receptor-regulated genes. Described in many cancers | Inhibits influenza virus replication |
| ATG16L2 | Involved in autophagy. Function is unknown | Might be a part of mucosal immune response |
| LTB4R | Is a receptor for leukotriene B4, a potent chemoattractant involved in inflammation and immune response | See main function. Regulates inflammatory response. |
| ARHGEF1 | Regulator of Rho-signalling | Regulates lung immune response and inflammation via T-cell activity. Associated with rare antibody deficiencies. |
| GAA | Essential for the degradation of glycogen in lysosomes |  |
| PTMS | Regulates resistance to certain opportunistic infections |  |
| MAP1S | Mediates aggregation of mitochondria resulting in cell death and genomic destruction. Plays a role in apoptosis | Regulates phagocytosis of bacteria and innate immunity signaling. |
| DLGAP4 | May play a role in the molecular organization of synapses and neuronal cell signalling |  |
| DPP9 | Acts as an inhibitor of caspase-1-dependent monocyte and macrophage pyroptosis | Regulates cytokine signalling, involved in multiple immunologic processes. Increased in COVID-19 patients. |
| OTUB1 | Regulates ubiquitin-dependent pathways, T-cell anergy (tumor immune escape) | Inhibits influenza virus development |
| MARK2 | An important regulator of cell polarity in epithelial and neuronal cells | Involved in innate responses regulation |
| DNM2 | Plays an important role in vesicular trafficking processes, in particular. Involved in cytokinesis. |  |
| C17orf62 |  |  |
| MGC52000 | Proposed to be implicated in T-cell proliferation and effector function. In dendritic cells involved in endosome-to-membrane recycling of major histocompatibility complex (MHC) class II |  |
| HK3 | Mediates the initial step of glycolysis by catalyzing phosphorylation of D-glucose to D-glucose 6-phosphate | Regulates immune cells infiltration in cancers |
| FLJ14107 | Unknown. Associated with long non-coding RNAs |  |
| KCTD13 | Might be involved in regulation of replication processes. |  |

**Table 5 B.**
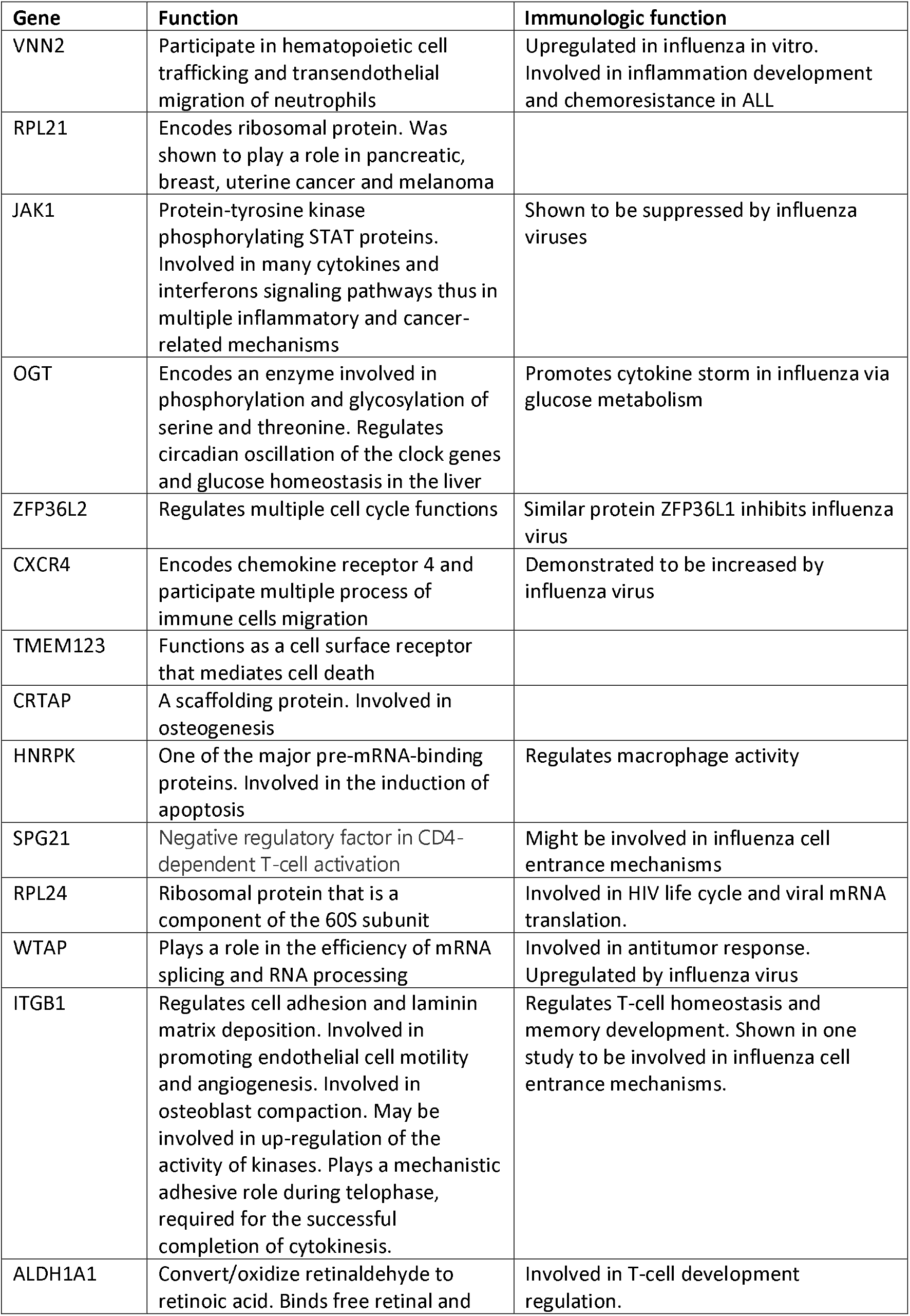
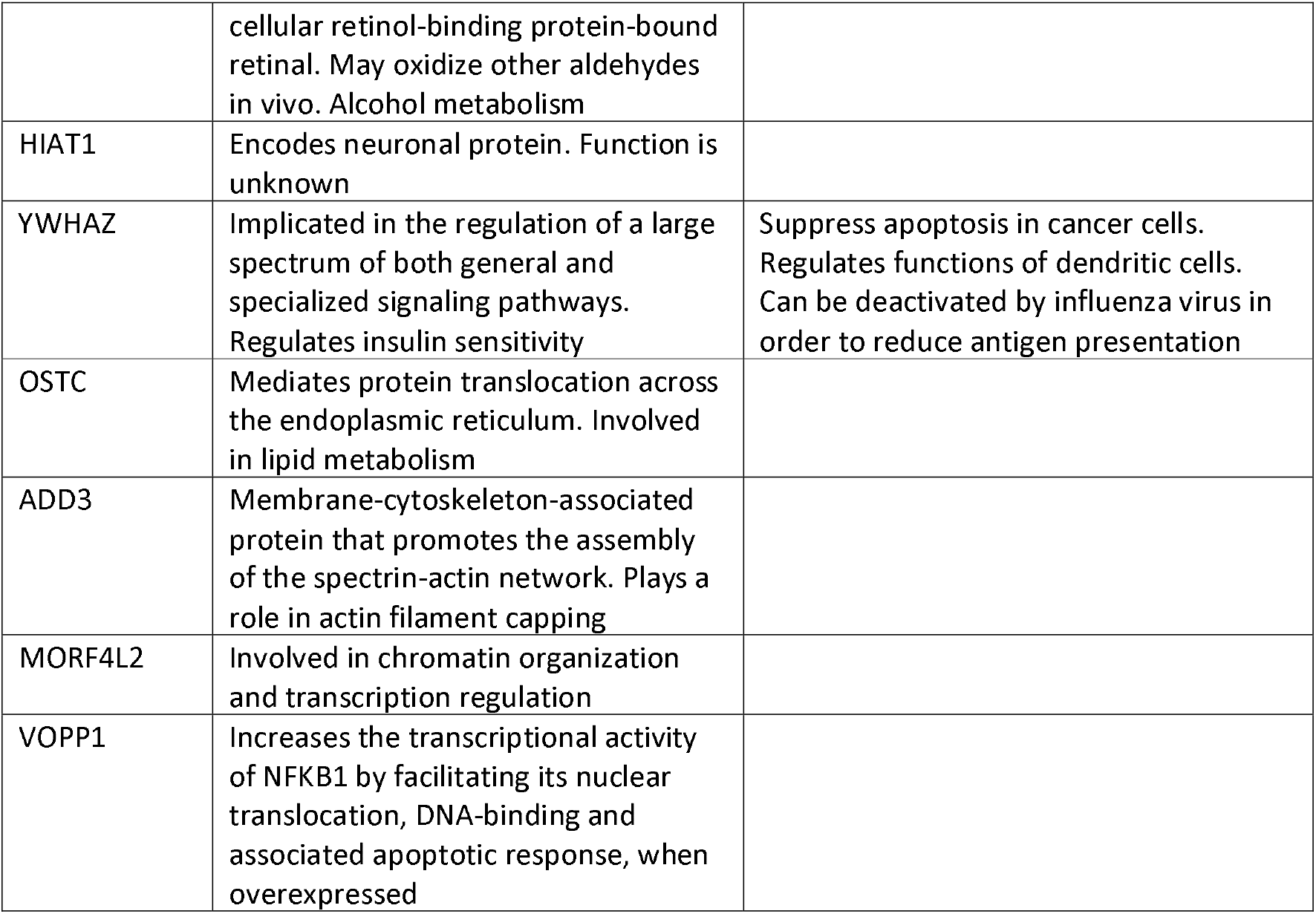
Gene names, function and immunological function of the 20 genes with largest negative values of *h*_*g*_.

So far in the analyses the only criteria for inclusion into the analyses was the p-values. In epidemiology this is always combined with a measure of relative risk. Here the fold change is the value of gene expression at the maximum compared to the average. A low fold change describes a very flat seasonal curve. Introducing a limit of log fold change => 0.1 or a fold change of 1.05, (5 % increase), reduced the number of genes significantly in both the Discovery and Replication datasets to 281, Table 6.

**Table 6.** Number of within-host genes (N=305) after running exclusion criteria based on log fold change for *c*_*g*_ ≥ 0.1.

| | $c_g$ discovery $\geq 0.1$ | $c_g$ discovery $<0.1$ |
| --- | --- | --- |
| $c_g$ replication $\geq 0.1$ | 281 | 9 |
| $c_g$ replication $<0.1$ | 14 | 1 |

A list of genes in a test for pneumonia infections like seasonal influenza and covid-19 was published recently (9). The test contains 36 genes as signature, Table 7. The number of genes available for analyses in our study depended on the different criteria in the preprocessing. More stringent criteria for detection limit decreased the number of genes. In a comparison of the genes in the test with the 1051 within-host list of our study, only one gene, GBP1, was in both lists.

**Table 7.**
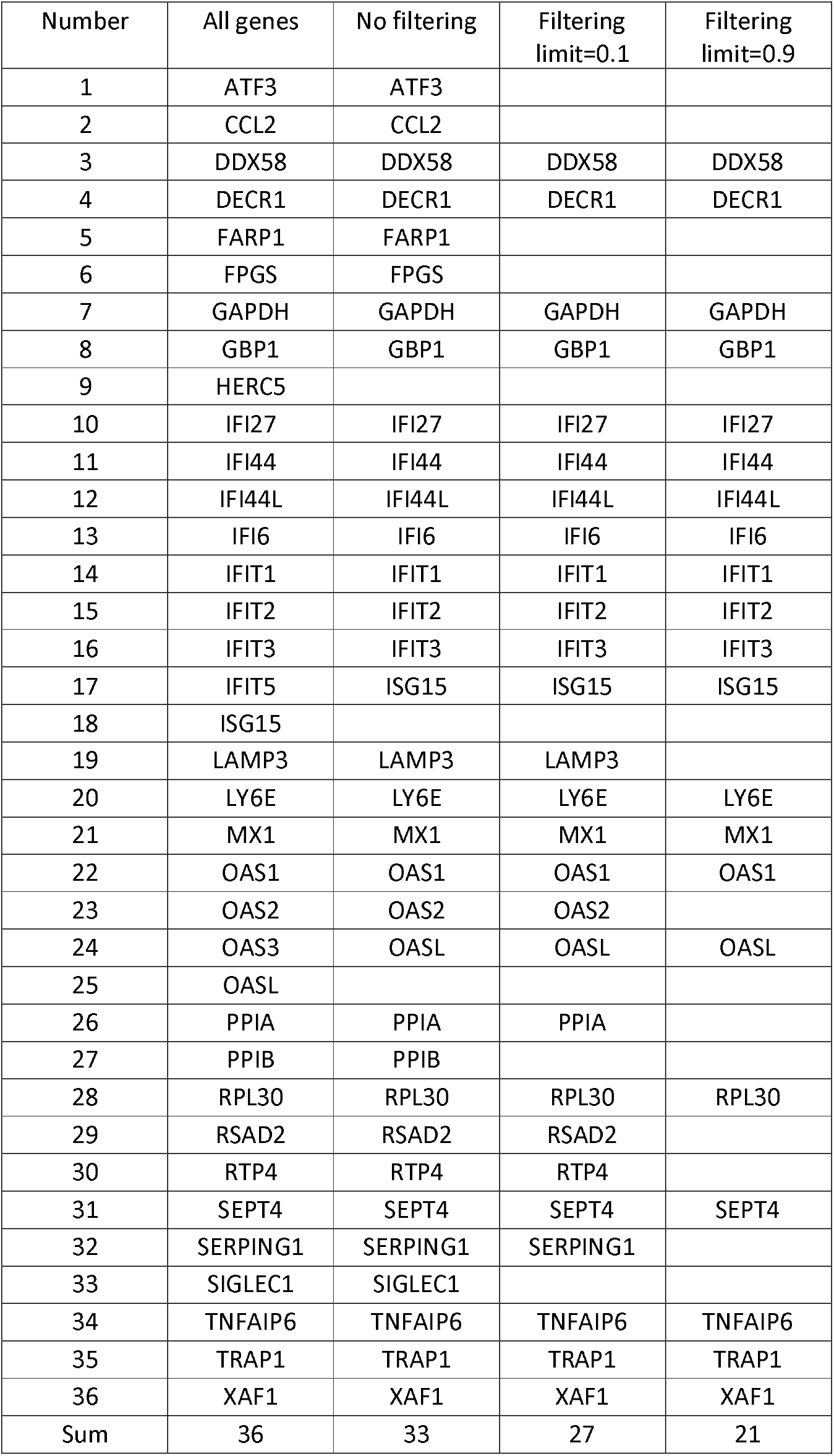
Gene expression test for influenza (McClain). Only one gene, GBP1, from the test was found in the 1051 gene list of potential within-host genes.

Lastly, in another analysis of covid-19 and severity of the disease (13) the genes OAS 1,2,3 were found to be protective. These genes were not significant in this study, Table 8.

**Table 8.** Significant expression of genes OAS1,2,3 in the discovery dataset dependent on preprocessing criteria (detectable yes/no and significant yes/no) and inclusion criteria of the within-host definition.

| Number of genes |  | OAS1 | OAS2 | OAS3 |
| --- | --- | --- | --- | --- |
| All genes | 25212 | yes | yes | yes |
| Detectable genes | 6133 | yes | no | no |
| Significant genes | 2942 | no | no | no |
| Within-host genes | 1051 | no | no | no |

## Discussion

This analysis has, for the first time to our knowledge, found a strong covariation between seasonal influenza and annual changes in gene expression for a subset of genes from immune cells in peripheral blood. The trajectories of these genes showed both a significant coefficient for annual seasonal changes and a flu determined term. The peaks of the gene expression level were observed to change concomitantly with the peaks of the epidemics independent of the month of the peak. The findings should have been externally validated in other studies, but a thorough search in Medline did not reveal similar studies (Box 1).

These findings support our operationalized within-host concept at a population level. Within-host seasonal immunity is a rather unspecified concept and the lack of knowledge on how the seasonality of these genes changes the immune system is challenging (1, 2, 5, 8). The applied definition of both a seasonal term and a flu term in the model could be one way of searching for genes participating in within-host seasonal immunity. From a total of 1000 genes with both terms significant the number was reduced to three hundred in the discovery - replication analyses.

These results indicate that during an influenza epidemic or pandemic changes occur on a population level for some genes. The regular seasonal pattern and the flu driven peaks indicates a relationship between these two. There were two major patterns of gene expression. In the first pattern, genes that have a maximum of seasonal expression in autumn were downregulated in winter, concomitantly with a decreased gene expression in many of the flu genes. The wave of changes in gene expression followed the flu intensities closely. In the second pattern, genes that were downregulated in late summer had a maximum seasonal effect in winter with mostly positive flu coefficients. The effect for the upregulated genes lasted beyond the flu. These findings could mirror two different biological responses to seasonal virus infections. The response of the within-host genes adapted to the timing of the virus epidemic was clearly seen for the year 2003, that demonstrated an early shift in response.

Moving the flu intensities up to 60 days before the observed flu intensities gave for many genes a better fit for the model with largest effect around three weeks earlier. This indicated that the changes in the coefficient for the flu term came before or at the onset of the flu epidemic in a within-host model. This could be the time from incubation till serious disease resulting in a sick leave. NIPH estimated a delay between infection and sick leave in the order of one week.

The changes in gene expression represented all infected women in the study population. During the epidemic as many as five percent of the total work force of around 2.5 million could have a sick leave for influenza in one week (Figure 1). Around 70% of women aged 16-74 were working. The participating women were middle aged and mainly aged 45-65 years at time of blood sampling. Using hypothesis testing, we were not able to show that the effect was due to a small group of infected people. That would have supported that the flu gene expression was due to the numbers of women with flu in the population.

From the end of 2021 a third wave of COVID-19 moves over the temperate regions of Europe and US (11). The seasonality of deaths due to the pandemic in Norway (Figure 1) demonstrates the three waves. The similarities with the years described here for seasonal influenza is striking (Figure 2). The covid-19 pandemic display new waves due to novel mutations, similar to seasonal influenza. The mutations give diseases with different symptoms and severity. Consequently, knowledge about the spread of seasonal influenza and a potential within-host seasonal immunity could have a transfer value to corona viruses (35). There are a few studies of covid-19 with results that implicit support of our view. In a study of blood gene expression profiles in patients admitted to emergency wards, strong transcriptional responses were found not only in COVID-19, but also for seasonal coronavirus, influenza, bacterial pneumonia, and healthy controls (9, 10). Proposed classifiers for the different serious infections and healthy controls showed auROC (mean area under the receiver operating characteristic curve) close to 1.0. However, of the 36 genes in the test only one was found among the 1051 genes named within-host genes here. Thus, the genes involved in the acute phase of serious disease did not confound the within-host genes. However, the healthy group of the control panel consisted of only 6 women, all 20 years. The lack of consistency between the within-host flu genes and the acute phase genes defined by the gene expression test could support our interpretations. Recently, the genes OAS 1,2,3 were found to be associated with improved survival of covid-19 (13). The genes did exist in our datasets, but only OAS1 passed the preprocessing limit. OAS1 did not reach significant expression level in the sine-cosine model.

The seasonal influenza epidemics showed for three of the four years of observation approximately the same pattern with infection rates increasing early in winter lasting for a few months before disappearing. In the year 2003, the epidemic came three months earlier. The expression of the within-host genes changed in the same direction. This indicated that neither light nor short daylight-induced vitamin D deficiency were important for the response (2). The seasonal patterns of genome-wide gene expression have been investigated in several studies (17, 18, 19). The same statistical model as used here were applied in other analyses with focus on summer and winter effects. The number of genes with significant seasonality varied among the studies. In a German study, 2311genes were upregulated in the summer and 2826 genes upregulated in the winter or 23% of all tested genes (17). They defined seasonal effect as positive relative effect in January, February, and December and negative in June, July and August. Another study from Australia (19) found less than two hundred significant seasonal genes. Lastly (18), a US study found 898 significant transcripts. These three studies were based on clinical studies of individuals with risk of prediabetes (36), children with increased risk of type I diabetes (37), and a study of familial melanoma (38). The models used by us were qualitatively very different and gave much more information opening for maximum response any date in the year. In the previous mentioned German study, the original analyses found a total of 5137 significant genes (17). This was reduced to 179 or 4% in an external validation study comparing the original findings with five samples from a multi-center study. Here the 1051 significant within-host genes were reduced to 305 or 30% in the replication analysis. One reason for the different results could be the design of the studies. By introducing a discovery – replication design the internal validity of the results in our study was verified. We are not aware of other studies looking at seasonal virus diseases that could be used for an external validation of our findings (Box 1).

It was recommended from the early beginning of gene expression analyses to use split sample methodology to control for weak associations and biases like technology sampling methods and preprocessing (39). Split sample analyses will have lower statistical power compared to using all samples in an explorative design. Here we used a split sample design with discovery and replication studies (25). The conditions for a good replication design are that the discovery (original) and the replication (confirmatory) populations are similar in terms of sex, age, ethnicity, and other important factors. In addition, the two studies should share identical laboratory analyses, data processing pipelines and analytical approaches. Thus, a replication study is methodologically different from studies of external validity. The replication analyses showed that many weak associations could not be reproduced. This reduced the problem of false positive results that can mislead later researchers. Both the peak months of the flu and the strength of the flu coefficient were significant in both datasets for one third of the genes.

Interpretation of significance solely on the p-values have given a very large number of significant genes in almost all analyses of seasonal gene expression, here close to 3 000. This could indicate that the strong parametrization of the sine – cosine model without replication design leads to many false positive results. For many decades epidemiology has relied more on relative risks than p-values alone (40). In functional genomics fold change is used as term for the difference between maximum amplitude and the average value for each gene. The introduction of a limit of the log fold change of 0.1 reduced the number of genes somewhat in a common analysis of the discovery and replication datasets. A log fold change less than 0.1 is equal to a relative risk of less than 5%. Such low risks can easily be confounded.

The study design has some limitations. The discovery and the replication samples were run on two different microarrays. The discovery set used Illumina H-6 with six blood samples on the chip, compared to the 12 blood samples on the Illumina Hu-12 in the replication. The shift was due to the sudden stop in the production of Illumina Hu-6. Hu-6 had approximately twice as many probes per gene as Hu-12 (Appendix Figure A). It might be that the reduction in probe numbers per gene reduced the sensibility of the microarray. Still a reasonable test-retest result was obtained. The original NOWAC study invited only women since it was based on hypotheses related to female reproduction, oral contraception, and hormonal replacement therapy. The age span included mainly women in their middle age. Due to the original design of building a biobank for functional genomics the study covered only four years. On the other hand, the sampling was independent of any knowledge of seasonal influenza.

A major issue in the interpretation of the findings is the lack of knowledge about many basic aspects of the antibody response to the influenza A virus (15). This makes it difficult to test specific hypotheses. Preliminary analyses of the gene expression did not support any specific hypothesis. This could be due to lack of knowledge of the biological relationship between host and viruses.

A major strength of this study was the completely random assignment for blood sampling regardless of place of living in Norway. Due to the extended sampling period, the design covered four years with different timing of the annual seasonal influenza. This population-based design increased the validity of the findings. As a national representative cohort, we could use national representative sickness data collected by the National Institute of Public Health. In the original case-control design, controls with cancer diagnosed before or during follow-up were excluded.

The discovery - replication design clearly demonstrated the need to discuss the current use of only p-values in studies of functional genomics due to the problem of false positive findings.

It is well accepted that ecological analyses should not be used for causal conclusions. Still, the identical fluctuations in gene expression and seasonal influenza ask for an explanation. This opens for creation of hypotheses. From an evolutionary point of view there could be several scenarios. First, if the seasonality of virus infections has lasted for thousands of years, individuals with an evolutionary adapted seasonal immune system could have better survival. On the other hand, if the immune system is seasonal for other reasons with a decreased effectiveness in the cold season as often proposed, then virus that operates in cold seasons would have an evolutionary better adaptation. Hypothetically, the within-host immune response could start in humans by the same signals of humidity and temperature that initiate the virus epidemics in the temporal regions, giving a within-host seasonal immunity in humans.

## Conclusion

To our knowledge, we have demonstrated for the first-time our concept of within-host seasonal immunity. For some genes we found both seasonality and influenza dependent changes in longitudinal analyses at a population level. This could represent important aspects for understanding potential new seasonal virus epidemics like covid-19. The results of our unique discovery – replication designed study have demonstrated the need for careful interpretations of results from functional genomic analyses.

## Data Availability

All data produced in the present study are available upon reasonable request to the authors.

## Supporting information

## Acknowledgement

We are thankful to all the women who donated blood samples to the NOWAC biobank. Bente Augdal and Merete Albertsen are thanked for their work on all infrastructure and administrative issues. We are grateful to Heidi Grønn for technical support. We are thankful for the valuable proof-reading of the English text by prof. emeritus Lise L. Håheim.

## Disclaimers

Microarray service was provided by the Genomics Core Facility, at the Norwegian University of Science and Technology, part of the Norwegian Microarray Consortium (NMC), a national technology platform supported by the functional genomics program FUGE of the Research Council of Norway.

## Author contributions

**Conceptualization:** Eiliv Lund, Lill-Tove Rasmussen Busund, Lars Holden

**Formal analysis:** Marit Holden, Lars Holden

**Funding acquisition:** Eiliv Lund, Lill-Tove Rasmussen Busund

**Methodology:** Lars Holden, Marit Holden, Igor Snapkov, Eiliv Lund

**Data handling:** Nikita Shvetsov

**Writing - original draft:** Eiliv Lund, Lars Holden

**Writing and discussions:** Eiliv Lund, Lars Holden, Lill-Tove Rasmussen Busund, Marit Holden, Igor Snapkov, Nikita Shvetsov

**Review and editing:** Eiliv Lund

## Funding

This study was supported by a grant given to EL from the European Research Council (ERC-AdG 232997 TICE), and a donation from Halfdan Jacobsen og frues legat (The Norwegian Cancer Society). The funders had no part in the study design, analyses or publication.

## Data available statement

Data will be available on request according to Norwegian health research law. Please contact corresponding author.

## Competing interest

The authors have declared that no competing interests exist.

### Box 1

Searched 19.01.2022

Database: Ovid MEDLINE(R) and In-Process, In-Data-Review & Other Non-Indexed Citations and Daily <1946 to January 18, 2022>

Search Strategy:

---------------------------------------------------------------------------------------------------------------------------

1. exp Seasons/ (114470)
2. seasonal variation*.ti,kw. (6975)
3. circannual rhythm*.ti,kw. (187)
4. 1 or 2 or 3 (115696)
5. exp Gene Expression/ (481411)
6. gene expression*.ti,kw. (115013)
7. transcriptomic*.ti,kw. (16226)
8. 5 or 6 or 7 (563996)
9. 4 and 8 (726)
10. exp Orthomyxoviridae/ (61015)
11. influenza virus*.ti,kw. (21372)
12. 10 or 11 (64103)
13. 9 and 12 (8)

Search for: 9 and 12

Results: 8

Six of the 8 were related to vaccines, one a clinical study and one cell study.

## Appendix figures

**Appendix Figure A.**
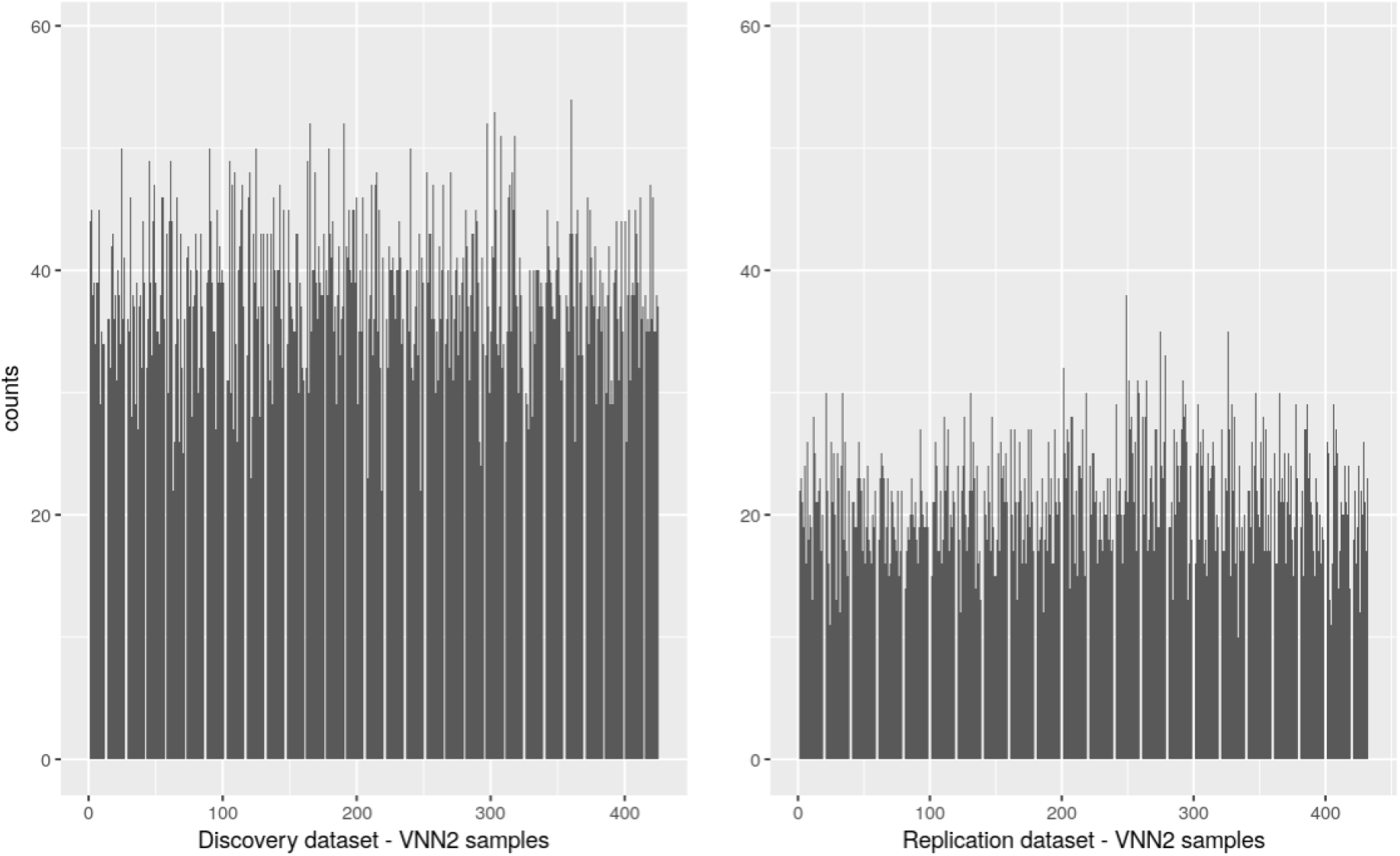
Plot of the distribution of the beads for the same gene VVN2 from Hu-6 (discovery dataset) and Hu-12 (replication dataset.

**Appendix Figure B.**
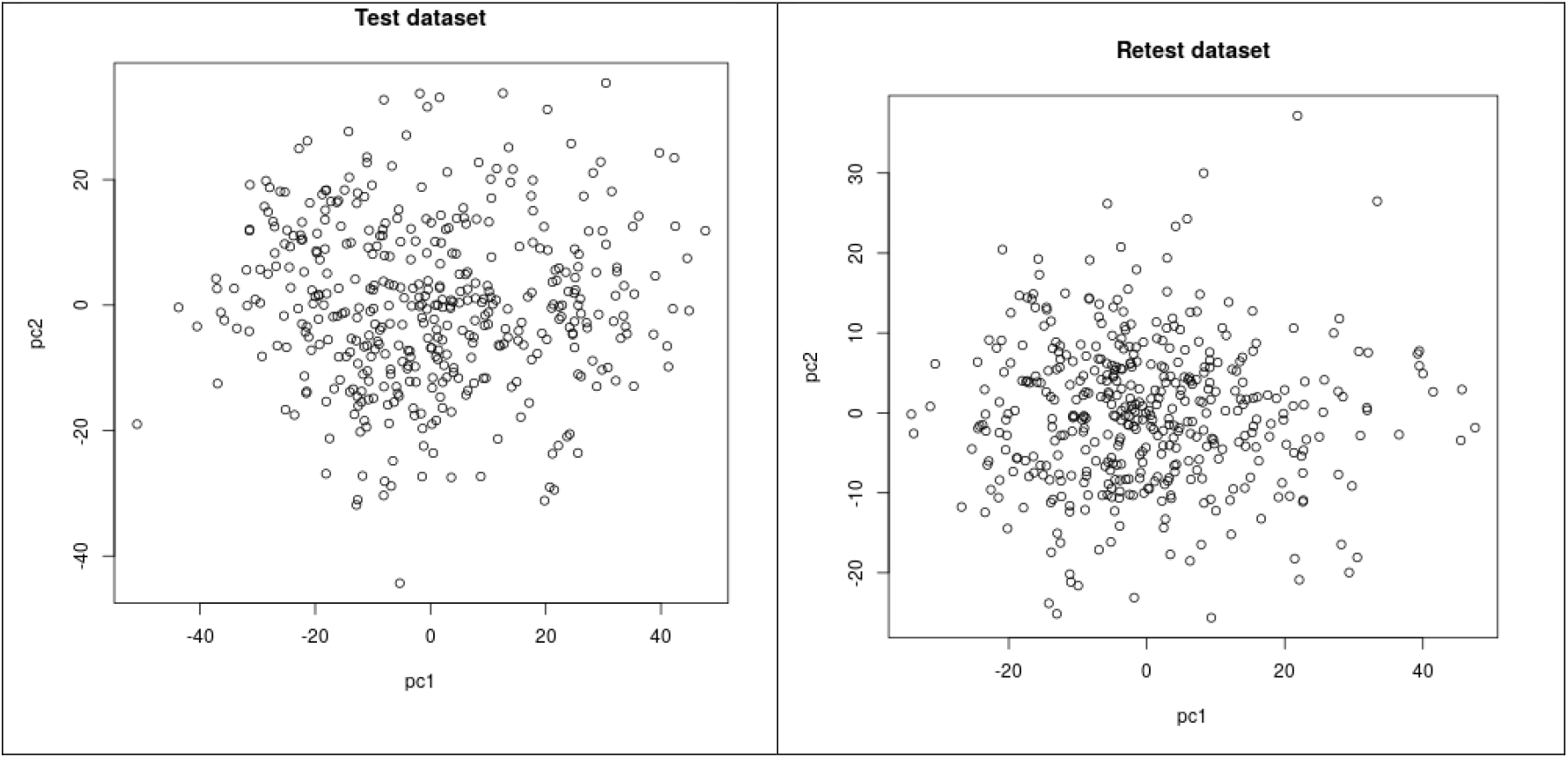
Plot of the first and second principal component of the discovery dataset, (left panel), 6118, genes, 425 samples after we have removed five outliers based on the PCA plots. Replication dataset (right panel) 6348 genes, 432 samples. The two datasets were pre-processed separately using a p-value for cut-off *p* = 0.01, and a present limit of 0.9.

## Notes

### Competing Interest Statement

The authors have declared no competing interest.

### Funding Statement

This study was supported by a grant given to Eiliv Lund from the European Research Council (ERC-AdG 232997 TICE), and a donation from Halfdan Jacobsen og frues legat (The Norwegian Cancer Society). The funders had no part in the study design, analyses or publication.

### Author Declarations

The NOWAC study was approved by the Norwegian Data Inspectorate and recommended by the Regional Ethical Committee of Northern Norway (REC North). The linkages of the NOWAC database to national registries such as the Cancer Registry of Norway and registries on death and emigration have also been approved. The women were informed about these linkages in the letter of invitation. Furthermore, the collection and storing of human biological material was approved by the REC North in accordance with the Norwegian Biobank Act. The linkages between Cancer Registry data and NOWAC study participants were performed at Statistics Norway, and the dataset was fully anonymized before it was made available to the authors. Information on breast cancer was used in this study to sample the random, matched controls. The Norwegian Data Protection Authority gave NOWAC exemption from the duty of confidentiality and permission to handle personal data (Datatilsynet, ref. 07/00030-2/cbr).

